# Tocilizumab and mortality in hospitalised patients with covid-19. A systematic review comparing randomised trials with observational studies

**DOI:** 10.1101/2021.04.23.21255815

**Authors:** Bélène Podmore, Nawab Qizilbash, Alessandra Lacetera, Itziar Ubillos, Kirsty Andresen, Ana Roncero Martín, Jara Majuelos-Melguizo, Ana Cuñado Moral, Marina Hinojosa Campos, Jeffrey K Aronson, Stuart Pocock

## Abstract

**Objective:** To summarise and compare evidence from randomised controlled trials and observational studies of the effect of tocilizumab on in-hospital mortality in patients with covid-19.

**Design:** Systematic review and meta-analysis.

**Data sources:** Searches conducted in Embase and PubMed from July 2020 until 1 March 2021.

**Study Selection:** Observational studies and randomised controlled trials (RCTs) assessing in-hospital mortality in patients receiving tocilizumab compared with standard care or placebo.

**Data extraction:** The primary outcome was in-hospital mortality at 30 days. The risk of bias in observational studies was assessed using the ROBINS-I tool. A fixed effect meta-analysis was used to combine relative risks, with random effects and risk of bias as a sensitivity analysis.

**Results:** Of 5,792 publications screened for inclusion, eight RCTs and 35 observational studies were identified. The RCTs showed an overall relative risk reduction in in-hospital mortality at 30 days of 0.86 (95% CI 0.78 to 0.96) with no statistically significant heterogeneity. 23 of the observational studies had a severe risk of bias, 10 of which did not adjust for potential confounders. The 10 observational studies with moderate risk of bias reported a larger reduction in mortality at 30-days (relative risk 0.72, 95% CI 0.64 to 0.81) but with significant heterogeneity (P<0.01).

**Conclusion:** This meta-analysis provides strong evidence from RCTs that tocilizumab reduces the risk of mortality in hospitalised covid-19 patients. Observational studies with moderate risk of bias exaggerated the benefits on mortality two-fold and showed heterogeneity. Collectively observational studies provide a less reliable evidence base for evaluating treatments for covid-19.

**Summary box:** *What is already known on this topic:*

- Early case reports suggested that tocilizumab might produce clinical and biochemical improvement in covid-19. This was followed by observational studies using retrospective data, largely supporting clinicians’ impressions of benefit.
- This led to wider use of tocilizumab, despite failure to show benefit on all-cause mortality from early underpowered randomised controlled trials (RCTs) in severe covid-19. The RECOVERY trial, the largest trial, has recently shown clear overall benefit in hospitalised patients with covid-19.

*What this study adds:*

- This meta-analysis provides strong evidence from RCTs that tocilizumab reduces the risk of mortality in hospitalised covid-19 patients.
- Observational studies with moderate risk of bias exaggerated the benefits on mortality by two-fold.
- Collectively observational studies provide a less reliable evidence base for evaluating treatments for covid-19.

## INTRODUCTION

Tocilizumab, currently licensed for treatment of rheumatoid arthritis, is a monoclonal antibody that inhibits interleukin-6 (IL-6) and is being used to treat patients with severe covid-19 (1).

IL-6 is a cytokine that is released by macrophages as part of the immune response to infection. Circulating IL-6 concentrations correlate with covid-19 severity (2). However, in severe covid-19 there is vascular inflammation and dysfunction, and IL-6 promotes endothelial dysfunction and impairs vascular permeability (3). Tocilizumab inhibits this inflammatory process.

Early case reports suggested that tocilizumab might produce clinical and biochemical improvement in covid-19 (4-6). This was followed by reports of observational studies using retrospective data, largely supporting clinicians’ impressions of benefit. This led to wider use of tocilizumab, despite failure to show benefit on all-cause mortality from early underpowered randomised controlled trials (RCTs) in severe covid-19. The RECOVERY trial has recently shown clear overall benefit in hospitalised patients with covid-19 of all degrees of severity, in addition to the benefit achieved with systematic corticosteroids (7).

We have therefore conducted a systematic review and meta-analysis of both randomised trials and observational studies of the effect of tocilizumab on in-hospital mortality.

## METHODS

### Real-time Systematic Review

This systematic review forms part of a real-time systematic review and meta-analysis that is updated regularly as new evidence becomes available. The protocol is registered on PROSPERO (Registration: CRD42020184089) and the EUPAS register (Registration: EUPAS35147).

### Search Strategy

A search of PubMed and Embase was conducted monthly from July 2020 until 1 March 2021, written in English, Spanish, French, and German, of treatment comparisons in hospitalised covid-19 patients and clinical outcomes. Search terms for treatment comparisons and clinical outcomes were combined with search terms for study design (randomised controlled trials and observational studies separately) (see Supplementary file 1). Where possible MeSH or index terms were used. We also searched the references in the retrieved papers for any additional relevant publications.

### Eligibility

All titles, abstracts, and selected full-text articles were reviewed for eligibility by five reviewers (KA, AC, AR, MH, JM). We included observational studies (either prospective or retrospective) and RCTs that reported the effect of tocilizumab on in-hospital mortality closest to 30 days in-patients with covid-19. Observational studies were eligible if they compared tocilizumab with standard care. Earlier publications that used the same data source over the same study period as a later publication were excluded as duplicates. RCTs were eligible if they compared tocilizumab against standard care or placebo. Studies that reported only mortality at 14 days or less were excluded.

### Data Extraction and Risk of bias assessment

For each RCT, data were extracted on study design (randomisation and blinding), comparator (placebo or standard care), the relative risk estimate, 95% confidence intervals, p values and analytic method. For each eligible observational study, information on study design, data source, population characteristics, outcome, analytical methods and covariate adjustments were extracted. A single measure was extracted from each study with adjusted measures in preference to unadjusted measures, where available. Where no measure of association was reported, the numbers of events were extracted. Data extraction was conducted by six reviewers (KA, JM, AC, AR, AL, MH) and any discrepancies were resolved by three separate senior reviewers (BP, SP, NQ).

The risk of bias was appraised using the Cochrane ROBINS-I (‘Risk Of Bias In Non-randomised Studies of Interventions’) tool for observational studies (8). Three reviewers (NQ, BP, IU) rated studies as being of low, moderate, serious or critical risk in each of the seven domains. Immortal time bias was assessed in the Bias due to Selection domain of the ROBINS-I tool. Any discrepancies in the assessment of the risk of bias were resolved with two senior reviewers (SP, NQ).

### Data synthesis and analysis

The first stage of data synthesis involved ensuring that a measure of association was available from each study. For studies in which no relative risk measure was reported, an unadjusted odds ratio was calculated. Owing to heterogeneity in the reporting of relative risks (rate ratio, hazard ratio, odds ratio), and the inclusion of both adjusted and unadjusted estimates from observational studies, the risk estimation methods could not be homogenised, and the reported relative risk estimates, were used as reported in each study.

The relative risk estimates from RCTs and observational studies were combined using both the inverse variance-weighted method for a fixed effect model and the DerSimonian-Laird random effect model. Heterogeneity was assessed using the I2 statistic and an interaction test p value, and corresponding forest plots were constructed. A sensitivity analysis to assess the effect of the risk of bias on the reported relative risk estimates was conducted. All analyses were conducted using R software (9).

### Patient and public Involvement

There was no involvement of patients or the public in the design, analysis, and interpretation of this study.

## RESULTS

The full search results are presented in the flow chart (see Figure 1). We have included 41 published comparative studies that evaluated the effect of tocilizumab on mortality in patients hospitalised with covid-19. These comprised eight randomised controlled trials (RCTs) (7, 10-16) and 33 observational studies (17-50). Study sizes ranged from 123 to 4,116 patients in RCTs and 33 to 3,924 patients in observational studies. The 41 studies came from 10 countries; the highest number (14 (34%)) came from the USA (see Supplementary file 2).

**Figure 1.**
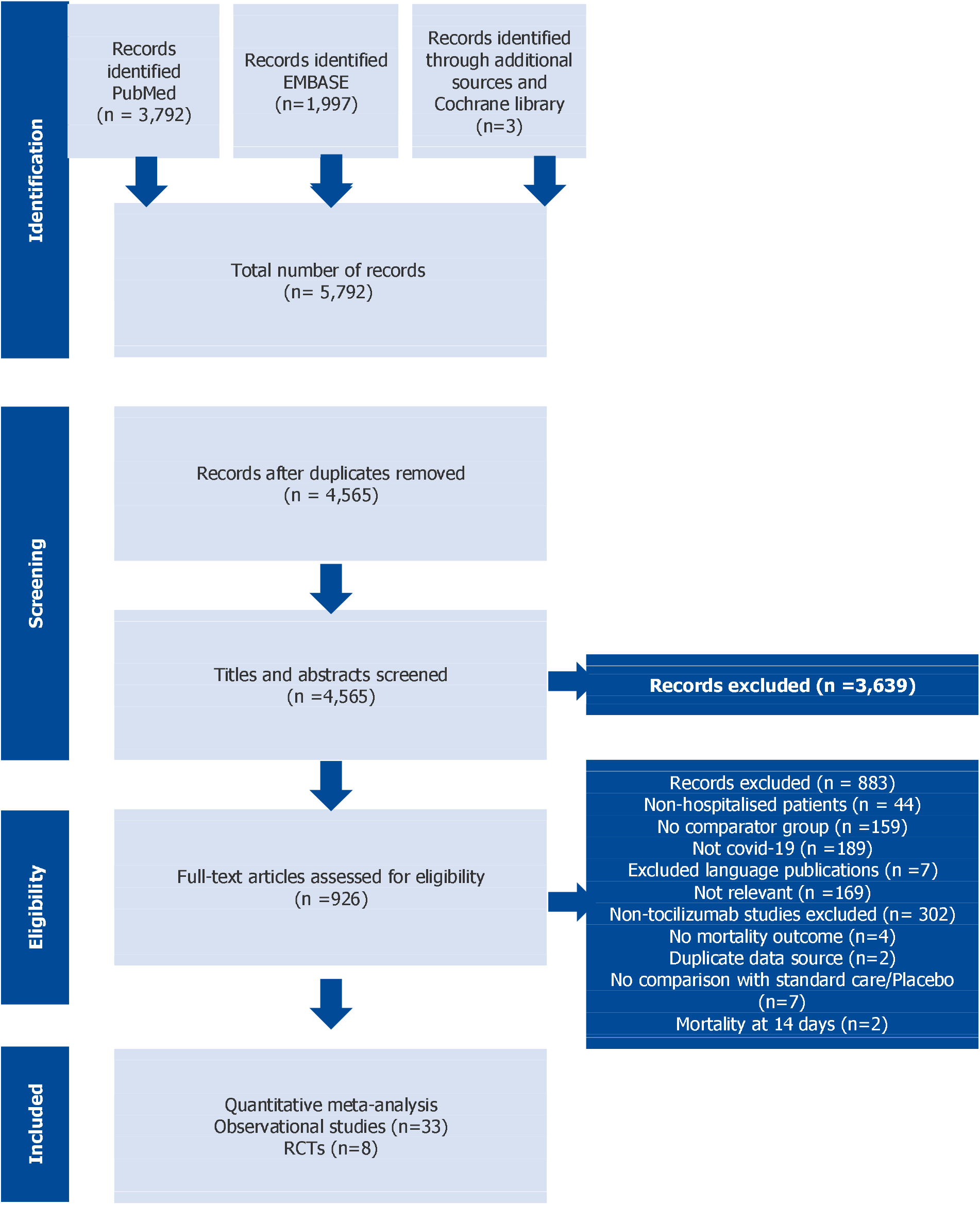
PRISMA flowchart

### The Randomised Evidence

Figure 2 presents a meta-analysis of the eight RCTs regarding the estimated relative risk effect of tocilizumab compared with standard care on 30-day mortality. In all cases, the dosage regimen was 8 mg/kg intravenously given once or twice (see Supplementary file 2). Using a fixed effect model, the combined estimate is a relative risk of 0.86 (95% CI 0.78 to 0.96, P=0.15).

**Figure 2.**
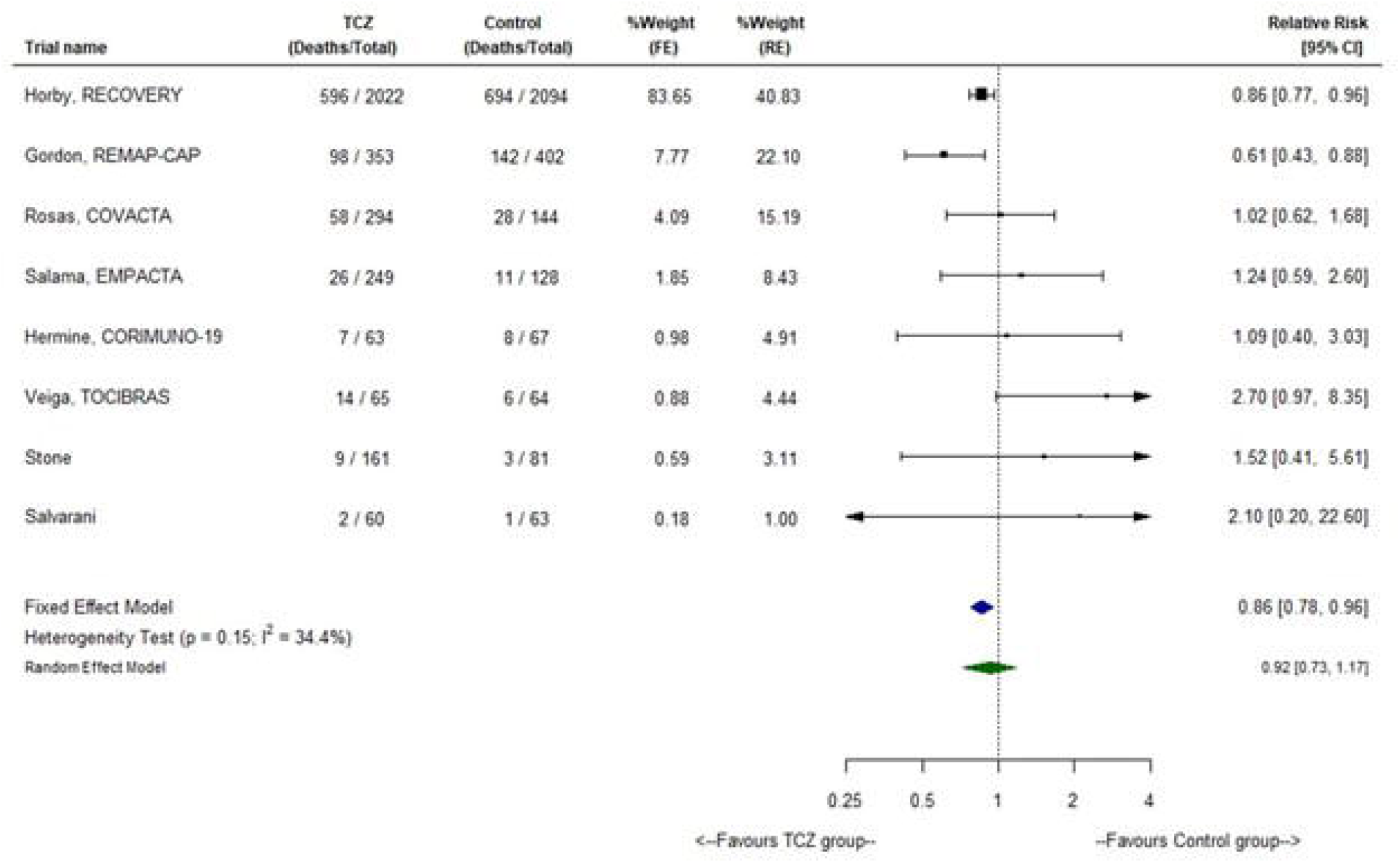
Fixed effects meta-analysis of RCTs reporting a relative risk for 30-day mortality

The RECOVERY trial is the largest, contributing 83.7% of the total weight. Hence, the meta-analysis produces a very similar treatment effect estimate to that in RECOVERY: relative risk 0.86 (95% CI 0.77 to 0.96). The next largest trial REMAP-CAP, weighted 7.8% and produced large treatment effect, relative risk 0.61 (95% CI 0.43 to 0.88). The other six smaller trials all had non-significant effect estimates, in the opposite direction.

The test for heterogeneity of effect sizes across trials was non-significant (interaction P=0.15). Nevertheless, this hint of apparent heterogeneity is sufficient to generate somewhat different results for a random-effects meta-analysis: the combined relative risk estimate of 0.92 comes nearer the null, with a wider 95% CI (0.73 to 1.17) and is non-significant. This arises because the random effect model gives increased weight to the six smaller studies (combined weight 37% compared with 8.8% in the fixed-effects model), and this pulls the overall estimate away from the highly positive RECOVERY result and increases the uncertainty.

While the absolute treatment benefit; the percentage reduction in mortality is of interest, it is hard to summarise. Since the mortality risk depends on the severity of the disease at the time of randomisation, it is plausible that the absolute treatment benefit will be more marked in patients with more severe disease. This could be explored in future subgroup analyses.

In RECOVERY, the percentage mortality reduction was 3.6% (95% CI 0.8 to 6.3)(7), while in REMAP-CAP (10) it was 7.3% (95% CI 0.95 to 13.2) (see supplementary file 4). Although the latter recruited more high-risk patients from intensive care units, the mortality rates in the control groups were similar (33% versus 35% respectively). We note that the five smallest RCTs all had much lower mortality rates (collectively 7.2%); it is, therefore, likely that they lacked the power to show a survival benefit of tocilizumab.

### Evidence from Observational Studies

The 33 observational studies comparing patients receiving tocilizumab against standard care are summarised in Figure 3. In all studies the dosage regimen was 4-8 mg/kg, to a maximum of 800 mg intravenously, given once or twice. We concentrate on the 23 studies that adjusted for potential confounders, separating the 10 other unadjusted studies as providing intrinsically unreliable evidence. Overall, the 23 adjusted observational studies produce a larger effect of tocilizumab on mortality than the RCTs. There is also significant heterogeneity among them (interaction P<0.01). Using a fixed effect model the overall relative risk is 0.72 (95% CI 0.65 to 0.80), whereas the random effect model estimate is 0.68 (95% CI 0.68 to 0.85).

**Figure 3.**
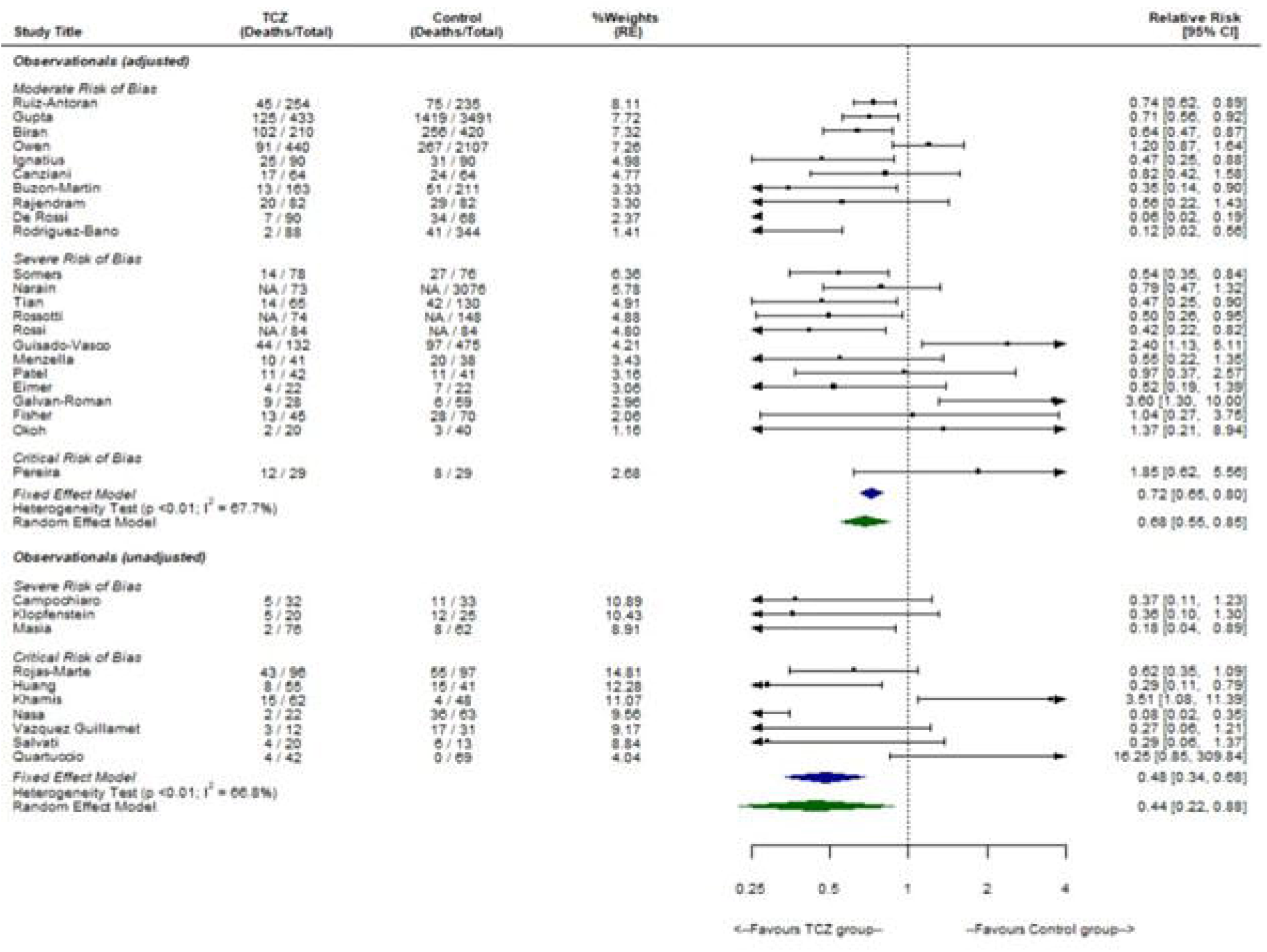
Fixed effects meta-analysis of observational studies reporting a relative risk of 30-day mortality by risk of bias.

Observational studies vary in their methodological quality. Of the 23 adjusted studies, 10 studies have a moderate risk of bias and 13 studies a severe or critical risk of bias. In Figure 4, we compared the treatment effect estimates for the RCTs with those for observational studies, split according to their risk of bias (moderate or severe). For the 10 observational studies with a moderate risk of bias the overall mortality relative risk from a fixed-effects model is 0.72 (95% CI 0.64 to 0.81), an apparently larger treatment effect than for the meta-analysis of RCTs (about twice as large a relative risk reduction).

**Figure 4.**
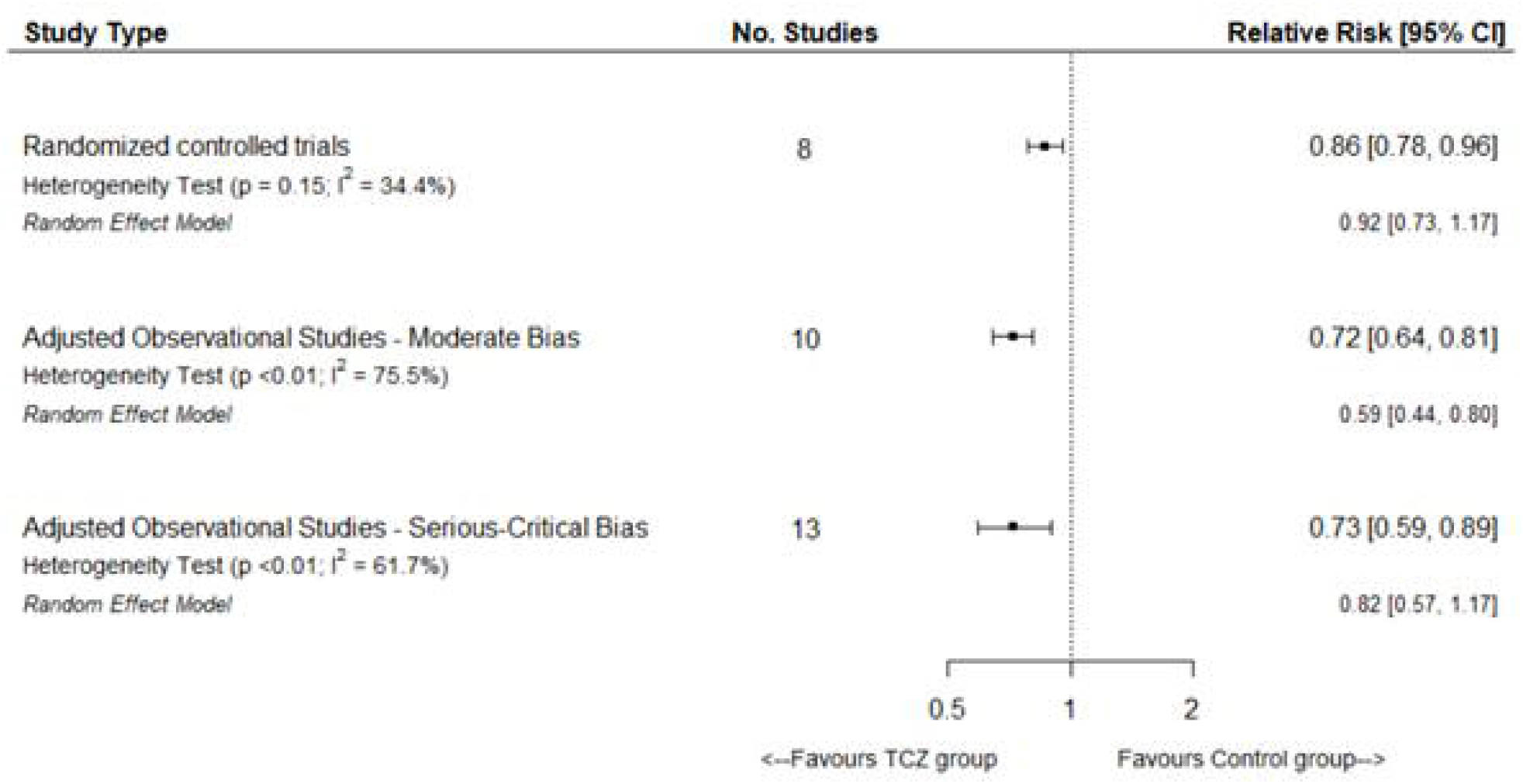
Forest plot of relative risk of 30-day mortality by study type and risk of bias

Four of the 10 studies dominate this overall estimate, with a combined weight of 66% in the fixed-effects meta-analysis. It is therefore worth exploring their methods. The largest cohort study (25) in patients admitted to 68 US intensive care units compared 433 patients who received tocilizumab within 2 days of admission, of whom 125 (29%) died with 3,491 patients who did not, of whom 1,419 (41%) died. Adjustment for over 20 potential confounders, using a propensity score with inverse probability weighting (IPW), resulted in a mortality hazard ratio of 0.71 (95% CI 0.56 to 0.92).

The second largest study (17), in patients admitted to 13 US intensive care units, included 210 patients who received tocilizumab, of whom 102 (49%) died. Of the 554 patients who did not receive tocilizumab, 420 were matched for propensity scores and 256 (61%) died. This involved adjustment for 13 potential confounders and correction for immortal time bias. The primary analysis yielded a mortality hazard ratio of 0.64 (95% CI 0.47 to 0.87).

The third study (50) was a retrospective cohort study of all patients with covid-19 in 17 Spanish hospitals. The 440 patients treated with tocilizumab had markedly higher unadjusted 28-day mortality than the other 2,107 patients (hazard ratio 2.35) but also had a poorer risk profile. After covariate adjustment for 22 factors (including corticosteroids) of which 13 were time-updated covariates, the hazard ratio became 1.20 (95% CI 0.87 to 1.64, P=0.26).

The fourth study (41) was in patients admitted to 18 tertiary hospitals in Spain with severe covid-19; 254 patients who received tocilizumab, of whom 45 (18%) died in hospital were compared with 235 patients who did not, of whom 75 (32%) died in hospital. Adjustment for over 20 potential confounders, using a propensity score with inverse probability weighting, resulted in a mortality hazard ratio of 0.74 (95% CI 0.62 to 0.89). It is puzzling that this study produces a substantially more precise treatment effect estimate (i.e., a narrower CI) than that of Gupta et al (25), even though it was around one-third the size. We suspect that this contradiction arises because the latter study correctly used a robust variance estimator to account for potential replication of patients induced by inverse probability, whereas the former did not.

## DISCUSSION

It is generally recognised that RCTs provide the highest quality of evidence on which to base therapeutic recommendations, while evidence from observational studies requires much more cautious interpretation. Hence, in interpreting this systematic review of the effect of tocilizumab on survival of patients with covid-19, it is appropriate that we first concentrate on the randomised evidence.

Overall, based on a fixed effect meta-analysis of eight RCTs, we see a 14% relative risk reduction in mortality with tocilizumab (95% CI 4-22%). This is very similar to the findings in the RECOVERY trial, which dominates the analysis, owing to its size.

We have also presented a random effect meta-analysis, since it is conventional to do so. It provides a weaker overall effect estimate, an 8% relative risk reduction with a wider 95% CI that includes no effect on mortality. However, this is likely to be a misleading analysis. There is no significant heterogeneity of effect across randomised trials (interaction P=0.15), yet the random effect model increases the weight given to the six smaller trials, none of which point in the direction of treatment benefit. This undue influence of small studies appears to dilute a treatment effect and generate increased uncertainty. There is a long-standing debate on the relative merits of fixed effect and random effect meta-analyses. In this case, we think that a random effect model is less trustworthy.

A key question is whether the overall survival benefit from tocilizumab relates to all hospitalised patients with covid-19 or if there are specific subgroups in whom the benefit is greater or absent. The RECOVERY trial reports that these benefits were seen in all patient subgroups, including those requiring oxygen, and those requiring mechanical ventilation in an intensive care unit (ICU)(7). The combination of tocilizumab and a systemic corticosteroid (e.g., dexamethasone) appears to reduce mortality to a greater extent. It is also plausible that the reduction in mortality due to tocilizumab is more marked in more severe disease, in which IL-6 release may be more marked. For instance, the second largest RCT, the REMAP-CAP trial, was in critically ill patients in an ICU and reported a 39% relative reduction in in-hospital mortality, although with a wide 95% CI (12-57%)(10). However, in the RECOVERY trial (7), patients requiring non-invasive ventilation and invasive mechanical ventilation (each subgroup having more deaths than in the REMAP-CAP trial) did not have larger relative reductions in in-hospital mortality than patients who did not require ventilatory support.

Interpretation of the evidence from observational studies presents more of a challenge. For the sake of completeness, we have included all 33 observational studies that evaluated the association between tocilizumab treatment and mortality (see Figure 3), but we feel it best to ignore the findings of most of them, owing to their unreliability. 10 studies did not adjust for confounders in their mortality analyses and a further 13 studies were classified as having a severe risk of bias. Reasons for such a poor rating include lack of adjustment for key covariates and bias in the selection of patients.

This leaves 10 observational studies classified as having a moderate risk of bias. Their combined data (2,093 patients given tocilizumab, of whom 460 died in hospital) amounted to a slightly lower mortality than in the RCTs. Overall, these 10 studies showed a stronger association between tocilizumab treatment and survival than the RCTs, with a relative reduction in mortality of 29% (95% CI 20-36%). The four largest were all retrospective cohort studies based on multiple hospitals, two in the USA and two in Spain. While the pooled 95% confidence interval in these studies at moderate risk of bias overlapped with the pooled estimate from the RCTs, it is noteworthy that only two observational studies had point estimates that fell within the 95% confidence interval of the pooled RCTs or the RECOVERY trial, the largest RCT.

The diversity of statistical methods across these studies is a challenge: propensity adjustment with IPW, propensity matching, and covariate adjustment were all used to account for potential confounders. Inevitably, one doubts whether any study has adequately corrected for the selection bias involved in the clinical decisions about who received tocilizumab and who did not. Unmeasured confounders may well play an important role. Hence, the extent to which one can trust the adjusted relative risk estimate in each observational study is open to debate, and the overall effect estimate across the 10 observational studies with moderate bias may have been overestimated two-fold (14% in RCTs versus 29% in observational studies).

Our systematic review has some limitations. We have only evaluated treatment effects on mortality, whereas other outcomes such as time to recovery and need for mechanical ventilation may have an important bearing on the overall benefit profile of tocilizumab and its cost-effectiveness by reducing the duration of illness. We believe that in-hospital mortality, as well as being the most important outcome, provides the least scope for bias in comparing RCTs and observational studies. We have concentrated on overall mortality in all patients, whereas there could be subgroups for whom the survival benefit is more (or less) marked, although subgroup analyses according to severity in the RECOVERY trial suggest that that is not the case.

The role of observational studies of treatments in covid-19, and more generally, is controversial. For tocilizumab, the pooled observational studies agree with the RCTs in the direction of benefit on mortality but exaggerated its magnitude two-fold. The large observational studies may seem to have been more informative motivation than the early underpowered RCTs which even when pooled showed no evidence of tocilizumab’s efficacy. We did not combine RCTs and observational studies with network meta-analysis, which may produce highly misleading results (51). The results of observational studies should be used mainly to generate hypotheses and to inform the design of RCTs and not as a basis for treating patients, except when RCTs are not reliable, as recently reported for the efficacy of prophylactic anticoagulation in covid-19 patients (52).

## CONCLUSION

This systematic review of all reported RCTs of tocilizumab versus standard care shows strong evidence that tocilizumab reduces mortality in severe covid-19. Observational studies of adequate methodological quality also provided evidence of efficacy, but the effect size was exaggerated two-fold. Collectively observational studies provide a less reliable evidence base for evaluating treatment for covid-19.

## Data Availability

Requests for data sharing should be sent to the corresponding author.

## Footnotes

### Contributors

NQ and BP conceived the study and wrote the protocol with JM and SP. KA and JM performed the literature searches. KA, JM, AC, AR, MH screened articles for inclusion. BP, NQ, IU conducted the risk of bias assessment. KA, JM, AC, AR, AL, MH extracted data for analysis. AL undertook the meta-analysis and produced forest plots and summary results, under supervision of BP, NQ and SP. BP, SP, NQ and JA wrote the first draft of the manuscript. All authors revised the manuscript and approved the final version. NQ is the guarantor for this work and accepts full responsibility for the conduct of the study, had access to the data, and controlled the decision to publish. The corresponding author (NQ) attests that all listed authors meet authorship criteria and that no others meeting the criteria have been omitted.

### Ethical approval

Not required

### Data sharing

Requests for data sharing should be sent to the corresponding author.

### Dissemination declaration

Dissemination to study participants or patient organisations is not applicable.

### Transparency statement

The author affirms that the manuscript is an honest, accurate, and transparent account of the study being reported; that no important aspects of the study have been omitted; and that any discrepancies from the study as originally planned (and, if relevant, registered) have been explained.

## Supplementary Files

S1 – Search strategy

S2 – Data Extraction Table

S3 – Risk of Bias assessment

S4 – RCT % Risk Difference

