## Supplementary file 1 for "Tocilizumab and mortality in hospitalised patients with covid-19. A systematic review comparing randomised trials with observational studies"

**EMBASE**

**.MP SEARCH**

1	(coronavirus or coronavirus infections or covid 2019 or SARS2 or SARS-CoV-2 or SARS-CoV-19 or severe acute respiratory syndrome coronavirus or coronavirus infection or severe acute respiratory pneumonia outbreak or novel cov or 2019ncov or sars cov2 or cov2 or ncov or covid-19 or covid19 or coronaviridae or corona virus).mp. [mp=title, abstract, heading word, drug trade name, original title, device manufacturer, drug manufacturer, device trade name, keyword, floating subheading word, candidate term word]

2	Coronavirus infection/

3	Coronavirinae/

4	1 or 2 or 3

5	(hydroxychloroquine or chloroquine or azithromycin or darunavir or cobicistat or ritonavir or remdesivir or ifn alfa or ifn beta or ivermectin or favipiravir or umifenovir or arbidol or nitazoxanide or oseltamivir or ribavirin or sarilumab or siltuximab or tocilizumab or il6 or il-6 or anakinra or baricitinib or tofacitinib or ruxolitinib or bevacizumab or nivolumab or pembrolizumab or pd1 or pd-1 or colchicine or eculizumab or imatinib or vafidemstat or regn3048 or regn3051 or convalescent plasma or apn01 or corticosteroids or nsaids or dexamethasone or treatment* or drug therap* or medication*).mp. [mp=title, abstract, heading word, drug trade name, original title, device manufacturer, drug manufacturer, device trade name, keyword, floating subheading word, candidate term word]

6	4 and 5

7	cohort analysis/

8	case control study/

9	cross-sectional study/

10	(epidemiologic stud* or case-control or casecontrol or cohort stud* or cohort-analysis or follow-up stud* or observational stud* or longitudinal or retrospective or cross-sectional).mp. [mp=title, abstract, heading word, drug trade name, original title, device manufacturer, drug manufacturer, device trade name, keyword, floating subheading word, candidate term word]

11	7 or 8 or 9 or 10

**12	6 and 11**

**13 limit 12 to dd=[YYYYMMDD – YYYYMMDD] (update each month)**

**PUBMED**

((coronavirus [MeSH]) OR ("coronavirus infections"[MeSH Terms]) OR (coronavirus [All Fields]) OR ("covid 2019") OR ("SARS2") OR ("SARS-CoV-2") OR ("SARS-CoV-19") OR ("severe acute respiratory syndrome coronavirus 2" [supplementary concept]) OR (coronavirus infection) OR ("severe acute respiratory" pneumonia outbreak) OR ("novel cov") OR (2019ncov) OR (sars cov2) OR (cov2) OR (ncov) OR (covid-19) OR (covid19) OR (coronaviridae) OR ("corona virus")) AND ("Hydroxychloroquine" OR "chloroquine" OR "azithromycin" OR "darunavir" OR "cobicistat" OR "ritonavir" OR "remdesivir" OR "IFN alfa" OR "IFN beta" OR "ivermectin" OR "favipiravir" OR "umifenovir" OR "arbidol" OR "nitazoxanide" OR "oseltamivir" OR "ribavirin" OR "sarilumab" OR "siltuximab" OR "tocilizumab" OR "IL6" OR "IL-6" OR "anakinra" OR "baricitinib" OR "tofacitinib" OR "ruxolitinib" OR "bevacizumab" OR "nivolumab" OR "pembrolizumab" OR "PD1" OR "PD-1" OR "colchicine" OR "eculizumab" OR "imatinib" OR "vafidemstat" OR "REGN3048" or "REGN3051" OR "convalescent plasma" OR "APN01" OR "Corticosteroids" OR "NSAIDs" OR "dexamethasone" OR "treatment*" OR "drug therap*" OR "medication*" ) AND ((epidemiologic studies [MeSH Terms]) OR "epidemiologic stud*" OR "epidemiology stud*" OR ("case-control studies" [MeSH Terms]) OR "case control" OR ("cohort studies" [MeSH Terms]) OR "cohort stud*" OR "cohort analysis" OR "follow up stud*" OR "observational stud*" OR "longitudinal" OR "retrospective" OR "cross-sectional")
