## Supplementary file 2 for "Tocilizumab and mortality in hospitalised patients with covid-19. A systematic review comparing randomised trials with observational studies"

**Table 1. Summary of RCT studies**

| **Trial name** | **Study design** | **Number of patients** | **Study setting** | **Inclusion criteria** | **Exclusion criteria** | **Tocilizumab dosage regimen** | **Outcome** | **Analytical method** |
| --- | --- | --- | --- | --- | --- | --- | --- | --- |
| Horby et al. - RECOVERY | Randomized controlled open-label platform trial | 2,022 TCZ  2,094 SC | UK | - Hospitalised - SARS-Cov-2 infection (clinically suspected or laboratory confirmed) | - Medical history that might, in the opinion of the attending clinician, put the patient at significant risk if they were to participate in the trial | IV TCZ (400-800 mg). A second dose may be given ≥ 12 and < 24 h later if, in the opinion of the attending clinician, the patient’s condition has not improved | 28-day mortality | Rate Ratio |
| Gordon et al. - REMAP-CAP | Multifactorial adaptive platform trial | 353 TCZ  402 SC | Global | Critically ill patients, aged >18 years, with suspected or confirmed covid-19, admitted to an ICU and receiving respiratory or cardiovascular organ support | - Presumption that death was imminent with lack of commitment to full support - Participated in REMAP-CAP within 90 days - Known hypersensitivity to TCZ or to any of their excipients - Pregnancy - Current documented bacterial infection - Patient with any of following laboratory results out of the ranges detailed below at screening should be discussed depending on the medication:   - ANC ≤ 1.0 × 10^9^/L   - Haemoglobin level: no limitation   - PLT < 50 G/L   - SGOT or SGPT > 5 N | IV TCZ (8 mg/kg, max. 800 mg). Additional dose could be administered 12-24 h later at the discretion of the treating clinician | Primary hospital survival | Bayesian cumulative logistic model (OR) |
| Rosas et al. - COVACTA | Randomized double-blind placebo-controlled trial | 294 TCZ  144 PBO | Global | Patients ≥ 18 years with severe covid-19 pneumonia confirmed by RT-PCR in any body fluid and evidenced by bilateral chest infiltrates on chest X-ray or CT-scan were enrolled. Eligible patients had blood O_2_ saturation ≤ 93% or PaO_2_/FiO_2_ < 300 mm Hg | Treating physician determined that:   - Death was imminent and inevitable within 24 h - Patient had active tuberculosis or bacterial, fungal, or viral infection other than SARS-CoV-2 | IV TCZ (8mg/kg infusion, max. 800 mg) | 28-day mortality | Weighted difference in % |
| Salama et al. - EMPACTA | Randomized double-blind placebo-controlled trial | 249 TCZ  128 PBO | Global | Patients ≥ 18 years (with no upper age limit) hospitalised with covid-19 pneumonia confirmed by a positive PCR test and radiographic imaging were eligible. Patients had a blood O_2_ saturation < 94% on ambient air | - Patient received CPAP - Patient received bilevel positive airway pressure - Patient received MV | IV TCZ (8 mg/kg, max. 800 mg). Additional dose could be administered 8-24 h after the first | 28-day mortality | Weighted difference in % |
| Hermine et al. – CORIMUNO-19 | Randomized cohort-embedded investigator-initiated multicentre open-label Bayesian trial | 63 TCZ  67 SC | France | Patients not requiring ICU at admission with moderate and severe pneumopathy according to the WHO Criteria of severity of covid pneumopathy.  Moderate cases:   - Showing fever and respiratory symptoms with radiological findings of pneumonia and requiring between 3 L/min and 5 L/min of O_2_ to maintain an O_2_ saturation (SaO_2_) of ≥ 97%   Severe cases meeting any of the following criteria:   - Respiratory distress (≥30 breaths/min) - O_2_ saturation of ≤93% at rest in ambient air - O_2_ saturation of ≤97% with O_2_ > 5 L/min - PaO_2_/FiO_2_ ≤300 mm Hg | Exclusion criteria included:   - Known hypersensitivity to TCZ - Pregnancy - Current documented bacterial infection - Patients with any of following laboratory results out of the ranges detailed below at screening: ANC 1.0 × 10^9^/L or less or PLT < 50 G/L | IV TCZ (8 mg/kg, max. 800 mg). One dose on day one, additional 400 mg dose on day 3 if O_2_ requirement was not decreased by >50% | 28-day survival | Age and centre adjusted multivariable Cox regression (HR) |
| Veiga et al. - TOCIBRAS | Randomized multicentre open-label parallel group superiority trial | 65 TCZ  64 SC | Brazil | Hospital in-patients aged ≥ 18 years with SARS-CoV-2 infection, confirmed by RT-PCR, and with symptoms >3 days. Eligible patients had severe or critical covid-19 with evidence of pulmonary infiltrates confirmed by chest CT scan or X-ray and were receiving supplemental O_2_ to maintain O_2_ saturation > 93% or had been receiving MV for < 24 h before analysis  In addition, at least two of the following criteria had to be met:   - D dimer > 2.74 nmol/L (> 1000 ng/mL) - CRP > 50 mg/L (> 5 mg/dL) - Ferritin >300 μg/L, or - LDH > ULN | Exclusion criteria included:   - Active uncontrolled infection - Raised AST or ALT levels > 5 times the ULN - Renal disease with an eGFR of < 30 mL/min/1.72 m^2^ | IV TCZ (8 mg/kg max. 800 mg). Single dose | 28-day hospital mortality | Logistic regression (OR) |
| Stone et al. –  BACC Bay TCZ Trial | Randomized double-blind placebo-controlled trial | 161 TCZ  81 PBO | USA | Individuals 19-85 years-old RT-PCR or IgM antibody assay confirmed SARS-CoV-2  Patients had >2 of the following signs:   - Fever (body temperature >38°C) within 72 h before enrolment - Pulmonary infiltrates - Need for supplemental O_2_ to maintain an O_2_ saturation > 92%   >1 of the following laboratory criteria also had to be fulfilled:   - CRP level > 50 mg/L - Ferritin level > 500 ng/mL - D-dimer level > 1000 ng/mL - LDH level > 250 U/L | - Receiving supplemental O_2_ at a rate > 10 L/min - If they had a recent history of treatment with biologic agents or small molecule immunosuppressive therapy - Receiving other immunosuppressive therapy that the investigator believed placed them at higher risk for an infection - Individuals had had diverticulitis | IV TCZ (8 mg/kg, max. 800 mg). Single dose | 28-day mortality | stratified Cox regression (HR) |
| Salvarani et al. | Randomized multicentre open-label trial | 60 TCZ  63 SC | Italy | Patients ≥ 18 years, with an instrumental diagnosis of covid-19 pneumonia confirmed by a positive RT-PCR for SARS-CoV-2 in a respiratory tract specimen  Other inclusion criteria were:   - Acute respiratory failure with a PaO_2_/FiO_2_ ratio between 200-300 mm Hg - Inflammatory phenotype defined by a temperature > 38 °C during the last 2 days, and/or serum CRP levels of ≥ 10mg/dL and/or CRP level ≥ 2 the admission measurement | - ICU admission - Known hypersensitivity to TCZ - Any condition preventing future admission to ICU (e.g. advanced age with multiple comorbidities) - Patients expressed will to avoid future intubation - Patients were not allowed to receive invasive or non-invasive MV | IV TCZ (8 mg/kg, max. 800 mg). First dose <8 h from randomization, second dose <12 h | 30-day mortality | Chi-square test in an asymptotic form and the relative risk with its bilateral 95% CI |

**Table 2. Summary of observational studies**

| **Study** | **Study design** | **Number**  **of patients** | **Study setting** | **Study population** | **Exclusion criteria** | **Tocilizumab dosage regimen** | **Comparator** | **Outcome** | **Analytical method** | **Covariate adjustments** |
| --- | --- | --- | --- | --- | --- | --- | --- | --- | --- | --- |
| **Adjusted** | | | | | | | | | | |
| Ruiz-Antorán et al. | Retrospective observational study | 254 TCZ  235 SC | Spain  (18 tertiary Spanish hospitals) | Patients ≥ 18 years with covid-19, confirmed by PCR on nasopharyngeal swab, who were consecutively admitted outside the ICU with documented pneumonia (by either imaging and/or the presence of rales/crackles on physical examination) with severe respiratory failure | - Patients < 18 years old - Those who died within 24 h after admission to hospital or after developing inclusion criteria | IV TCZ | Standard care | In-hospital mortality | IPTW-adjusted regression (HR) | Inverse probability weighting based on propensity score matching based on age, gender, HT, neurological exploration, diabetes mellitus, WHO ordinal scale, time from symptoms, confirmed infection, lymphocytes, neutrophils, PLT, prothrombin activation, temperature, LDH, and baseline medication use of ACE-inhibitors, LPV/r, HCQ, CCT, IFN, NSAID, moxifloxacin, remdesivir, and AZT |
| Gupta et al. | Retrospective observational study | 433 TCZ  3,491 SC | USA  (STOP-covid study 68 hospitals across USA) | Patients aged ≥ 18 years with laboratory-confirmed covid-19 admitted to an ICU directly attributable to covid-19 | - Enrolment in a RCT involving TCZ or other IL-6 antagonists - Hospitalisation for ≥ 1 week before ICU admission - Liver disfunction (AST/ ALT > 500 U/L) - Receipt of an IL-6 antagonist other than TCZ during the first 2 days of ICU admission - Receipt of TCZ before ICU admission | TCZ | Standard care | Mortality | Inverse probability weighted Cox regression (HR) | Inverse probability weighted using age, sex, race, ethnicity, BMI, HT, diabetes, CAD, congestive HF, current tobacco use, active cancer, home medications (statin, ACE inhibitor, ARB-2), days from symptom onset to ICU admission (≤3 vs >3), severity-of-illness covariates assessed on ICU admission and concurrent therapies received on ICU admission |
| Biran et al. | Retrospective observational study | 210 TCZ  420 SC | USA  (13 hospitals in Hackensack Meridian Health Network) | Patients ≥ 18 years with laboratory-confirmed covid-19 who needed support in the ICU  Patients receiving TCZ for chronic rheumatological conditions were not excluded | - Pregnancy - Individuals participating in a clinical therapeutic trial | IV TCZ (400 mg). Second dose was permitted at worsening oxygenation and before mechanical ventilation | Standard care | Hospital-related mortality | Multivariable Cox regression (HR) | Age, gender, diabetes, COPD or asthma, HT, cancer, renal failure, obesity, oxygenation < 94%, qSOFA score, use of steroids, CRP > 15 mg/dL, intubation or MV support and time to TCZ treatment after admission |
| Owen et al. | Retrospective observational study | 440 TCZ  2,107 SC | Spain  (17 Grupo HM Hospitals) | Admitted to any of the participating hospitals with a diagnosis of covid POSITIVE or covid PENDING | None reported | Tocilizumab | Standard care | 28-day all-cause in hospital mortality | Multivariable Fine & Gray Model | Age, sex, Confirmed covid-19 diagnosis, supplemental O_2_, treatment with steroids, temperature (c), heart rate (bpm), SaO_2_ (%), SBP, DBP, CCI, Prior MI, congestive HF, PVD, cerebrovascular disease, dementia, pulmonary disease, renal disease, HT, diabetes, cancer, liver disease, prior stroke, ischemic heart disease, obesity, ALT, AST, creatinine, CRP, D-dimer, eosinophils, glucose, LDH, lymphocytes, monocytes, neutrophils, PLT count, potassium, sodium, urea, WBC count |
| Ignatius et al. | Retrospective observational study | 90 TCZ  90 SC | USA  (John Hopkins Health System, Washington DC) | Patients > 18 years with confirmed covid-19, hospitalised  The intervention group was patients who received TCZ for off-label treatment of covid-19, and the comparator arm was drawn from patients with covid-19 who did not receive TCZ | Patients were excluded if they were < 18 years old or if they died or were discharged within 24 hours after hospitalisation | IV TCZ (usually 8 mg/ kg, range 6–8 mg/kg, max. 800 mg). Single dose | Standard care (HCQ, AZT, CCT, heparin, remdesivir) | 28-day mortality | Inverse-probability weighted Cox regression (HR) | Age, sex, race, BMI, CCI, SpO_2_/FiO2, respiratory rate, temperature, SBP, DBP, pulse, O_2_ supplementation device, code status, CRP, WBC, ALC, Hgb, albumin, ALT, GFR, D-dimer, ferritin, and IL-6 |
| Canziani et al. | Retrospective observational study | 64 TCZ  64 SC | Italy  (Two general hospitals in Milan and Bergamo) | Adult patients with covid-19 in need of respiratory support Criteria for receiving TCZ:   - Clinical worsening in the previous 24 h with increasing need for O_2_ or ventilatory support - Absence of clinical or biochemical signs of an active bacterial infection - Elevated CRP - A higher risk for mortality at blood tests including lymphocyte count, ferritin, creatine kinase, ALT, and D-dimer | - Late intubation (> 24h) | IV TCZ (8 mg/kg). Second dose 24 h later if no clinical worsening had occurred between infusions | Standard care (SB ENX, direct antivirals including LPV/r, DRV + CBT, HCQ) | 30-day Mortality | Multivariable Cox regression (HR) | Matching variable (matched according to the respiratory support) and multivariable adjustment (variables were selected if the rate of missing values was very low (<5%) and proved significant in the univariable Cox analysis (P value < 0.1) |
| Buzón-Martín et al. | Retrospective observational study | 163 TCZ  211 SC | Spain (University hospital of Burgos, Burgos) | Patients ≥ 18 years and admitted presenting covid-19 related respiratory insufficiency upon clinical and blood gas parameters | - Patients who died within 48 h of admission - Those testing positive but asymptomatic were excluded | TCZ | Standard care (respiratory support, LPV/r, AZT, HCQ, ENX, IFN 1-β and methylprednisolone) | Mortality | Multivariate Cox regression (HR) | Adjustment not listed |
| Rajendram et al. | Retrospective observational study | 82 TCZ  82 SC | USA  (Ten hospitals within the Cleveland Clinic Enterprise) | Patients with RT-PCR confirmed SARS-CoV-2 and admitted to the ICU at the time of TCZ administration | - Received additional doses of TCZ more than 48 h after the initial dose - Received TCZ through an RCT | IV TCZ (4–8 mg/kg max. dose 400 mg). Single dose - additional doses discouraged | Standard care | 28-day mortality | Multivariable logistic regression (OR) | Propensity score matching based on ICU admission source, max. CRP, SOFA score at ICU admission, vasopressor use, age, race, weight, and the use of MV during hospital admission and multivariate adjustment (not listed) |
| De Rossi et al. | Retrospective observational study | 90 TCZ  68 SC | Italy (Montichiari Hospital) | - Confirmed covid-19 infection by a positive RT-PCR collected on a nasopharyngeal swab - Bilateral pulmonary interstitial opacities on chest imaging that were not fully explained by congestive HF or other forms of volume overload - Respiratory failure ≥ 1 of the following conditions: - Respiratory rate ≥ 30 breaths/min - SpO_2_ ≤ 93% while breathing ambient air - PaO_2_/FiO_2_ ≤ 300 mm Hg | - Presence of a critical respiratory syndrome that requires IMV or MV at hospital admission - Presence of severe clinical conditions as revealed by ALT 5x ULN - Neutrophils <500 mmc and/or PLT <50.000 mmc | IV TCZ (400 mg) or SB TCZ (324 mg) | Standard care (HCQ, LPV/r) | Death | Multivariable Cox regression (HR) | Age, gender, diabetes, HT, heart diseases, serum CRP, respiratory support needed at hospital admission, and time elapsed from symptoms onset to hospital admission |
| Rodríguez-Baño et al. | Retrospective observational study | 88 TCZ  344 SC | Spain  (60 Spanish hospitals) | Presenting on a specific date (day 0) with > 1 clinical and 1 laboratory criterion suggestive of a hyperinflammatory state:   - Temperature ≥ 38°C - Increase in O_2_ support required to achieve O_2_ saturation > 92% - Laboratory criteria were ferritin > 2000 ng/mL or increase >1000 ng/mL since admission, D-dimers > 1500 mg mL (or doubled in 24 h), and IL6 > 50 pg/mL | - Being under MV at day 0 - Occurrence of the primary endpoint in ≤ 2 day after day 0 - Written decision to avoid any escalation in medical treatment before day 0 - Previous use of systemic CCT, TCZ, other immunomodulatory drugs or immunoglobulins - Treatment with immunomodulatory drugs other than CCT or TCZ, or with immunoglobulins during the first 48 h after day 0 | TCZ | Standard care | 21-day mortality | Inverse probability weighted Cox regression (HR) | Inverse probability weighting calculated using propensity score matching based on age, gender, ethnicity, comorbidities (cardiac disease, HT, chronic pulmonary disease, chronic renal disease, liver cirrhosis, malignancy, diabetes mellitus, obesity, HIV infection), laboratory data (lymphocytes, LDH, ALT, ferritin, D-dimers, IL-6), previous treatments, radiographic findings, 7-point scale and type of O_2_ requirement |
| Somers et al. | Retrospective observational study | 78 TCZ  76 SC | USA  (Covid-19 Rapid Response Registry) | - Admitted for severe covid-19 pneumonia, had a RT-PCR positive SARS-CoV-2 test, and required IMV - Individualized decisions on TCZ usage were made by the attending infectious diseases physician | - < 16 years - Intubated for conditions unrelated to covid-19 - Enrolled in an RCT for sarilumab | IV TCZ (8 mg/kg max. 800 mg). Single dose - additional doses discouraged | Standard care | 28-day Mortality | Inverse probability weighted Cox regression (HR) | Inverse probability weighting based propensity score matching (age, congestive HF, chronic pulmonary disease, chronic renal disease, therapeutic anticoagulation, ferritin, LDH, and AST) |
| Narain et al. | Retrospective observational study | 73 TCZ  3,076 SC | USA  (12 hospitals and emergency departments within Northwell Health system) | - Patients > 18 year with covid-19 positivity as determined by PCR testing of nasopharyngeal swabs - Meeting CCS criteria of ferritin > 700 ng/mL or CRP > 30 mg/dL or LDH > 300 U/L | - Received any prespecified immunomodulatory drug (steroids, anakinra, TCZ) before the patient met the inclusion criteria | TCZ | Standard care (i.e. AZN, HCQ, colchicine and ascorbic acid) | In-hospital mortality | Multivariable Cox proportional hazards (HR) | Age, sex, race or ethnicity, smoking history, insurance status, treated in a tertiary vs community medical centre, chronic lung disease, CV disease, HT, diabetes, renal disease, haemodialysis, liver disease, cancer, autoimmune disease, CCI, BMI, CRP, ferritin, D-dimer, LDH, haemoglobin, PLT, serum sodium, serum transaminases, neutrophil-to-lymphocyte ratio, use of IMV and vasopressor use within 24 h |
| Tian et al. | Retrospective observational study | 65 TCZ  130 SC | China  (Tongji Hospital, Wuhan Pulmonary Hospital, and Renmin Hospital of Wuhan University, Wuhan) | Patients ≥ 18 years with covid-19 admitted to hospitals  Criteria for administering TCZ:   - Diagnosis of covid-19 confirmed upon RT-PCR positivity for SARS-CoV-2 - Patients with extensive lung lesions - Severe cases who also show an increased level of IL-6 in laboratory testing | - Incomplete medical records (e.g., transfer to any other hospital) - Evidence of concomitant bacterial infection, and pregnancy | IV TCZ (400 mg max. 800 mg). Second dose within 12 h in case of fever. Third dose 24 h apart based on clinician response | Standard care (i.e., antiviral, antibiotics, immunoglobulin therapy, CCT) | In-hospital death | Multivariable Cox regression (HR) | Propensity score matching based on age, sex, and comorbidities (HT, diabetes, tumour, coronary heart disease, chronic obstructive pulmonary disease, cerebral infarction, liver cirrhosis, hepatitis, and tuberculosis) and adjustment using time to death, controlling for treatment group and potential confounders, including age, gender, and comorbidities |
| Rossotti et al. | Retrospective observational study | 74 TCZ  148 SC | Italy  (ASST Grande Ospedale Metropolitano Niguarda, Milan) | Patients ≥ 18 years with a RT-PCR SARS-CoV-2 with a diagnosis of severe or critical covid-19 according to the Chinese Guidelines for the management of covid-19 Criteria for administering TCZ:   - CT scan findings of severe, bilateral interstitial pneumonia - Presence of an active inflammatory status alternatively defined by abnormal CRP levels (> 1 mg/dL), IL-6 > 40 pg/mL, d -dimer > 1.5 mcg/mL, or ferritin > 500 ng/mL) | - ALT value > 5 x ULN; neutrophil cell count < 500 cell/mmc; PLT count < 50,000 cell/mmc - An active bacterial infection or a complicated intestinal diverticulitis - A positive pregnancy test - A positive HBsAg status - Any concomitant disease not defined as “under control” | IV TCZ (8 mg/kg infused max. dose of 800 mg). Second dose after 12 h in case of fever persistence | Standard care | In-hospital mortality | Cox regression (HR) | Matching based on age, sex, severity of disease, P/F, CCI, and length of time between symptoms onset and hospital admittance |
| Rossi et al. | Retrospective observational study | 84 TCZ  84 SC | France (Primary care centre regional hospital) | - Severe covid-19 pneumonia defined as pulse SpO_2_ ≤ 96% despite O_2_ support ≥ 6 L/min with O_2_ mask, for > 6 h - TCZ was administered at discretion of the attending physician | - Patients with invasive MV (i.e., intubated) - Patients in the critical care medicine department | IV TCZ (400 mg). Single dose | Standard care | 28-day Mortality | Cox regression (HR) | Inverse probability weighting based on propensity score matching based on age, sex, smoking status, history of CAD, stroke, HF or PAD, HT, chronic kidney disease with estimated GFR < 60 mL/min/1.73 m^2^, cancer, long-term CCT treatment, use of antibiotics, antivirals, CCT or baricitinib after admission, SpO_2_/FiO_2_ ratio at admission, time between admission and inclusion, and SpO_2_/FiO_2_ ratio and CRP at inclusion |
| Guisado-Vasco et al. | Retrospective observational study | 132 TCZ  475 SC | Spain (Hospital Universitario Quirón salud Madrid) | Those admitted to the hospital with covid-19 pneumonia:   - Clinical criteria:   - Pneumonia confirmed by chest imaging   - SaO_2_ ≤ 94% while breathing ambient air or ratio of the PaO_2_/FiO_2_ ≤ 300 mm Hg or SaO_2_/FiO_2_ of 235 and 315 mm Hg - Microbiological criteria:   - PCR confirmed SARS-CoV-2 by nasopharyngeal swab during admission   - After an initial negative PCR result, but a typical clinical scenario of SARS-CoV-2 infection   - Distinguished clinical picture even without conducting a PCR assay, according to local epidemiology | - Pregnancy or breast-feeding - < 18 years old - Known allergy or hypersensitivity to any drug in the protocol, advanced dementia, vital prognosis < 6 months, chronic renal insufficiency with a filtration rate < 25 ml/min/1.73m2, untreated hepatitis B or C infection, known severe liver disease, previous uncontrolled arterial HT, prolonged QTc interval at triage - Any concomitant medication that contraindicated any of the selected drugs in the protocol - Patients who were only under supportive care owing to their severe condition | TCZ | Standard care (HCQ, AZT, LPV/r, DRV–CBT and LMWH for prophylaxis) | In-hospital mortality | Binary logistic regression model (OR) | Multivariate adjustment (adjustment variables not listed) |
| Menzella et al. | Retrospective observational study | 41 TCZ  38 SC | Italy (IRCCS of Reggio Emilia, Reggio Emilia) | Patients with SARS-CoV-2 infection confirmed by a positive RT-PCR assay in a respiratory tract specimen and clinical and radiological findings compatible with covid-19 severe pneumonia. The criteria for administering TCZ were strictly based on drug availability | None reported | IV TCZ (8 mg/kg max. 800 mg) by two consecutive infusions 12 h apart.  SC TCZ (162 mg) ranging from 2 to 4 doses depending on drug availability and body weight due to a temporary unavailability of IV formulation | Standard care (antimicrobial and/or immunomodulatory therapy containing LPV/r, HCQ, AZT, interferon, remdesivir, methylprednisolone) | In-hospital mortality | Multivariable Cox proportional hazards (HR) | Age, sex |
| Patel et al. | Retrospective observational study | 42 TCZ  41 SC | USA (Swedish Medical Centre, Washington) | Patients ≥ 18 years old hospitalised for covid-19 and treated with TCZ and a comparative matched cohort that did not receive TCZ | Patients enrolled in RCTs of TCZ | TCZ | Standard care | Mortality | Not reported | Matching based on exact WHO score at hospital administration and TCZ administration, day of TCZ administration and age |
| Eimer et al. | Retrospective observational study | 22 TCZ  22 SC | Sweden (Karolinska University Hospital Huddinge, Stockholm) | Patients > 18 years with confirmed SARS-CoV-2 infection admitted to the ICU for severe ARDS  Criteria for receiving TCZ (at discretion of the physician):   - Rising O_2_ requirements with > 5 L/min on O_2_ mask to maintain SpO_2_ at 94% - ≥ 7 days from symptom onset - Hyperinflammation characterized by > 1 of the following: - CRP > 100 mg/L or doubled in the last 24 h - LDH > 8 µkat/L - IL-6 > 40 ng/L - D-Dimer > 2mg/L - Rising, high sensitivity troponin T > 15 ng/L - Ferritin > 500 µg/L - No contraindication to TCZ - AST/ALT at > 5 times of the upper limit of normal, - Neutropenia with < 500 cells/mm3, - Thrombocytopenia < 50 cells/mm3)   Patients were eligible as controls if they were admitted to ICU with covid-19 and ARDS did not fulfil the TCZ treatment criteria | Patients with positive PCR for SARS-CoV-2 admitted for a primary diagnosis other than ARDS | IV TCZ (8 mg/kg). Single dose | Standard care | 30-day mortality | Not reported (HR) | Propensity score matched using age, diabetes, HT, obesity, D-dimer, IL-6, troponin T and PaO_2_/FiO_2_ ratio |
| Galván-Román et al. | Retrospective observational study | 28 TCZ  59 SC | Spain (Hospital Universitario La Princesa, Madrid) | Patients with confirmed detection of SARS-CoV-2, baseline IL-6 serum level measurement and admitted to hospital with severe to critical covid-19  Criteria for administering TCZ:   - Interstitial pneumonia with severe respiratory failure (score = 2) - Rapid respiratory worsening requiring MV/IMV (score ≥ 3 on the covid respiratory severity scale) - Presence of extrapulmonary organ failure (shock or score ≥ 3 on the SOFA scale) - Severe systemic inflammatory response (IL-6 (> 40 pg/mL increased levels of D-dimer (> 1500 ng / mL); progressively increasing D-dimer) - Patients who, according to their baseline clinical condition, would be IMV subsidiary | IL-6 > 40 pg / ml | IV TCZ (8 mg/kg max. 800 mg). Second dose after 12h | Standard care | Mortality | Multivariable Cox Regression (HR) | Total lymphocyte count, D-dimer, LDH, PaO2/FiO2, COPD, obesity, HT, CRP, and IL-6 |
| Fisher et al. | Retrospective observational study | 45 TCZ  70 SC | USA  (Stony Brook University Hospital, New York) | Covid-19 pneumonia confirmed by nasal swab and required IMV in any ICU during their hospitalisation  The criteria for receiving TCZ (at discretion of primary healthcare provider):  Respiratory support in the form of high-flow nasal cannula or higher | None reported | IV TCZ (400 mg). Second dose after 24 h if there was a perceived lack of response to the initial dose | Standard care | 30-day Mortality | Multivariate logistic regression (OR) | Age, sex, BMI, SOFA score, CCI, baseline IL-6, CRP, ferritin, and CCT therapy |
| Okoh et al. | Retrospective observational study | 20 TCZ  40 SC | USA  (Newark Beth Israel Medical Centre, New Jersey) | Patients > 18-years with laboratory-confirmed SARS-CoV-2 with full clinical data who had completed their hospitalisation Criteria for administering TCZ:   - Decrease in WBC count, serum haemoglobin and PLT count - Elevation in baseline inflammatory markers: serum ferritin, LDH, procalcitonin, ESR and CRP - Patients who did not receive TCZ in addition to the standard care could not be treated with TCZ because:   - admitted to the general medical ward and managed as mild cases   - Existing bacterial infection   - chronic immunosuppression   - Unavailability of TCZ at the time of request | None reported | IV TCZ (8 mg/kg max. 800 mg). Second dose ≥ 12 h in patients who remain febrile within 24 h of initial dose | Standard care (HCQ, LPV/r, favipiravir) | In-hospital mortality | Chi-squared  Fishers Exact test | Propensity score matching based on age, gender, race, BMI, laboratory markers such as white cell count, haemoglobin, platelets, ferritin, CRP, LDH, ESR, procalcitonin, albumin and medications (HCQ, antibiotics, steroid) |
| Pereira et al. | Retrospective observational study | 29 TCZ  29 SC | USA (Columbia University Irving Medical Centre Hospital, New York) | Patients >18 years with solid organ transplant ≥ 90 days of potential observation  Criteria for administering TCZ:   - Patients with > 7 days of symptoms - Progressive respiratory distress - Rising levels of inflammatory markers including CRP, ferritin, or IL-6 | None reported | IV TCZ (4-8 mg/kg max. 800 mg). Additional doses of TCZ when the primary team deemed the initial response to be insufficient | Standard care | Mortality | Not reported | Matching based on age (> or < 60 years), HT, CKD, and receipt of high dose CCT |
| **Unadjusted** | | | | | | | | | | |
| Campochiaro et al. | Retrospective observational study | 32 TCZ  33 SC | Italy  (San Raffaele Hospital, Milan) | Criteria for receiving TCZ:   - Diagnosis of covid-19 confirmed upon positive RT-PCR for SARS-CoV-2 on nasopharyngeal swab - Hyper-inflammation (CRP, ≥ 100 mg/L, normal values <6 mg/L) or ferritin (≥ 900 ng/mL), in the presence LDH > 220 U/L) - Severe respiratory involvement defined by typical radiological findings at chest X-ray and/or CT scan, in the presence of an SaO_2_ ≤92% while breathing ambient air or PaO_2_/FiO_2_ ≤300 mm Hg - Patients admitted to hospital before or after the time period of TCZ availability who retrospectively fulfilled eligibility criteria for TCZ treatment were used as a comparison group | - Evidence of concomitant bacterial infection - History of diverticular disease - Neutropenia < 1500 × 10^9^ cells/L - Concomitant use of other immunosuppressive biologic drugs - Baseline elevation of AST/ALT levels > 5x ULN range - No concomitant CCT therapy | IV TCZ (400 mg). Second dose (400 mg) after 24 h in case of respiratory worsening | Standard care (HCQ, LPV/r, ceftriaxone, AZT, anti-coagulation prophylaxis with SB ENX) | 28-day Mortality | Two tailed Fisher's exact | No |
| Klopfenstein et al. | Retrospective observational study | 20 TCZ  25 SC | France  (Nord Franche-Comté Hospital, Trévenans) | Adult patients who received TCZ for confirmed COVID-19 by SARS-CoV-2 RT-PCR or diagnosis confirmed during the tocilizumab multidisciplinary team meeting.  Criteria for administering TCZ:   - No contraindication to TCZ - Confirmed covid-19 with SARS-CoV-2 RT-PCR (or high suspicion of covid-19 with obvious clinical, biological, and imaging data and without differential diagnosis despite a negative SARS-CoV-2 RT-PCR) - Failure of standard care - Time to symptom onset > 7 days - O_2_ therapy ≥ 5 l/min, - > 25% of lung damages on CT scan - ≥ 2 parameters of inflammation or biological markers of mortality (with a high level) such as ferritin, CRP, D-dimers, lymphopenia, and LDH - The standard care included patients receiving standard treatment but without TCZ   Control group: adult patients with confirmed COVID19 by SARS-CoV-2 RT-PCR receiving standard treatment but without tocilizumab | Control group:   - Patients with treatment not routinely administered in the hospital (remdesivir and immunoglobulins) - Patients with moderate disease - Those hospitalised < 48 h and/or who did not receive the standard treatment and/or O_2_ therapy) | TCZ. Single dose or two doses | Standard care | Death | Chi-squared or Fishers exact test | No |
| Masiá et al. | Retrospective observational study | 76 TCZ  62 SC | Spain (University Hospital of Elche, Elche) | All patients admitted confirmed or suspected covid-19 Criteria for administering TCZ:   - CURB-65 ≥ 2, O_2_ saturation < 93% - Respiratory frequency > 30 per min Chest X-ray with bilateral multilobar infiltrates - D-dimer ≥ 0.7 µg/L; IL-6 ≥ 40, pg/mL; lymphocyte count <800 × 10^9^/L; ferritin ≥ 700 µg/L; fibrinogen > 700 mg/dl; CRP > 25 mg/L |  | IV TCZ (600 mg if ≥75 kg or 400 mg if <75kg). Second dose after 24 h if persistence of fever; no improvement in tachypnoea; no improvement in SaO_2_ ≥ 5%; no decrease in CRP > 25%; radiological progression) | Standard care (antimicrobial and/or immunomodulatory therapy containing LPV/r, HQC, AZT, IFN-β-1b or remdesivir ± methylprednisolone | Death | Chi-squared or Fishers exact test | No |
| Vazquez Guillamet et al. | Retrospective observational study | 12 TCZ  31 SC | USA  (Washington University-Barnes Jewish Hospital, Washington) | Consecutive patients infected with SARS-CoV-2 requiring MV.  Criteria for administering TCZ was at discretion of treating physician. | None reported | IV TCZ (8 mg/kg). Second dose 12–24 hours later, if the clinical circumstances persisted | Standard care | 30-day mortality | Chi squared/Fishers exact test | No |
| Rojas-Marte et al. | Retrospective observational study | 96 TCZ  97 SC | USA (Maimonides Medical Centre, New York) | Adult patients hospitalised with severe to critical SARS-CoV-2 infection   - Severe disease: defined as requiring O_2_ supplementation via face mask up to 10 L/min to maintain an O_2_ saturation of ≥ 95% - Very severe disease: defined by requiring a non-rebreather mask or HFNC to maintain an O_2_ saturation of ≥ 95% - Critical disease: defined by the need for intubation and MV   Control group: patients required to be on supplemental O_2_ that matched the treatment group | - Died < 24h of admission - Included in clinical trials with other biologic agents or convalescent plasma | IV TCZ. Single dose | Standard care | Mortality | Chi-squared  or Fishers exact test | No |
| Huang et al. | Retrospective observational study | 55 TCZ  41 SC | USA  (Cedars Sinai Medical Centre, California) | Patients admitted for a covid-19-related admission with diagnosis confirmed by a positive nasopharyngeal RT-PCR test for SARS-CoV-2  Criteria for administering TCZ:   - Signs of respiratory compromise consisting of tachypnoea, dyspnoea OR - Peripheral capillary SpO_2_ < 90% on at least 4 L of O_2_ - Increasing oxygen requirements over 24 h, PLUS - > 2 of the following predictors for severe disease:   - IL-6 > 10 pg/mL   - CRP > 35 mg/L   - Ferritin > 500 ng/mL   - D-dimer > 1 mcg/L   - Neutrophil-Lymphocyte Ratio > 4   - LDH > 200 U/L   - Increased troponin in a patient without known cardiac disease | - Patients administered investigational IL-6 antagonist, clazakizumab - Non- covid related death | IV TCZ (400mg). Single dose | Standard care (HCQ, AZT, remdesivir, dexamethasone) | Mortality | Chi-squared/ Fisher’s exact test | No |
| Khamis et al. | Retrospective observational study | 62 TCZ  48 SC | Oman  (Tertiary care hospital, Muscat) | Patients hospitalised with confirmed or imminent respiratory failure and any one of the following conditions:   - ARDS; Severe pneumonia; Pneumonia; Critical respiratory condition requiring HFNC, IMV, MV, or rapidly increasing O_2_ requirement; Sepsis; Septic shock; MODS   Criteria for administering TCZ:   - Confirmed critical respiratory condition, rapidly increasing O_2_ requirements or severe covid-19 pneumonia as evidenced wit chest X-ray or CT scan and >1 of the following:   - Blood O_2_ saturation ≤ 93%   - PaO_2_/FiO_2_ < 300 mm Hg, - And:   - Established presence of hyperinflammation as per serial monitoring of serum ferritin, CRP, fibrinogen, d-dimer, LDH and IL-6     - Ferritin > 300 µg/L (or surrogate) and doubling within 24 h     - Ferritin > 600 µg/L at presentation and LDH > 250 U/L     - Elevated d-dimer (> 1 µg/mL)     - IL-6 > 80 pg/mL | - Coexistent infection other than covid-19 - History of severe allergic reactions to mAb - Long-term oral medication of anti-rejection drugs or immunoregulatory drugs, - Neutrophils < 500/μL or platelets < 50 × 10^9^ - Active diverticulitis, IBD, or another symptomatic GI tract condition that might predispose patients to bowel perforation; - Severe haematological, renal or liver function impairment (ALT/AST ratio > 5 ULN) - Active tuberculosis or other active infection | IV TCZ (4–8 mg/kg) followed by an additional dose after 12 h if no clinical response without exceeding a total of 800 mg | Standard care (including HCQ, LPV/r and IV steroids and O_2_ therapy) | Death | Chi-squared or Fisher’s exact test | No |
| Nasa et al. | Retrospective observational study | 22 TCZ  63 SC | United Arab Emirates  (2 centres in Dubai) | Severe and critical covid-19 patients who developed severe or critical CRS and no contraindications:   - Severe cases (New organ dysfunction: liver test dysfunction, acute kidney injury, sepsis: IVF for resuscitation, low dose vasopressor, supplemental O_2_ (HFNC HFNBM, FiO_2_ ≥ 40%, NIV) - Critical (Life-threatening, MV, high dose vasopressors) | None reported | IV TCZ (8 mg/kg max. 800 mg). Two divided doses 12 h apart | Standard care | 28-day Mortality | Chi-squared  Fishers Exact test | No |
| Salvati et al. | Retrospective observational study | 20 TCZ  13 SC | Italy  (Careggi University Hospital, Florence) | Adult patients admitted for covid-19 pneumonia | - Patients with evidence of bacterial sepsis - ANC < 500/mm^3^, thrombocytopenia (< 50,000 PLT/mm^3^), liver impairment (ALT > 2.5 times ULN), medical history positive for GI perforation - Known hypersensitivity to TCZ | IV TCZ (8 mg/kg max. 800 mg). Two doses twice 12−24 h | Standard care (Supplemental O_2_ therapy, LMWH, HCQ and LPV/r (or darunavir/cobicistat) | 28-day Mortality | Not reported | No |
| Quartuccio et al. | Retrospective observational study | 42 TCZ  69 SC | Italy  (Single centre hospital) | Patients with covid-19 pneumonia who provided oral or written consent | None reported | IV TCZ (8 mg/kg). Single dose | Standard care (IV methylprednisolone at 1 mg/kg/day) | Mortality | Chi-squared  Fishers Exact test | No |

*ACE inhibitor: angiotensin-converting enzyme inhibitor; ALT: alanine aminotransferase levels; ANC: absolute neutrophil count; ARB-2: angiotensin 2 receptor blocker; ARDS: acute respiratory distress syndrome; AST: aspartate aminotransferase; AZT: azathioprine; CAD: coronary artery disease; CBT: cobicistat; CCI: Charlson Morbidity Index; CCS: coronavirus disease 2019 cytokine storm; CCT: corticosteroids; COPD: chronic pulmonary obstructive disorder; CPAP: continuous positive airway pressure; CRP- c-reactive protein; CT: computed tomography; CV: cardiovascular; DBP: diastolic blood pressure; DRV: darunavir; GFR: glomerular filtration rate; ENX: enoxaparin; ESR: estimated sedimentation rate; GI: gastrointestinal; Hgb: haemoglobin; HCQ: hydroxychloroquine; HF: heart failure; HFNBM: high flow (> 10 L) Nonrebreathing mask; HFNC: high flow nasal canula; HR: hazard ratio; HT: hypertension; IBD: inflammatory bowel disease; ICU: intensive care unit; IFN: interferon; IL-6: interleukin-6; IMV: invasive mechanical ventilation; IV: intravenous; LDH: lactate dehydrogenase; LMWH: low molecular weight heparin; LPV/r: lopinavir + ritonavir; mAb: monoclonal antibodies; MI: myocardial infarction; MODS: Multiple Organ Dysfunction Syndrome; MV: mechanical ventilation; NIV: non-invasive ventilation; NSAID: non-steroidal anti-inflammatory drugs; O_2_: oxygen; OR: odds ratio; PAD: peripheral artery disease; PaO_2_/FiO_2_: ratio of arterial oxygen partial pressure to fractional inspired oxygen; PBO: placebo; PLT: platelets; PVD: peripheral vascular disease; qSOFA: quick sequential organ failure assessment; RT-PCR: reverse-transcriptase polymerase chain reaction; SaO_2_: oxygen saturation; SB: subcutaneous; SBP: systolic blood pressure; SC: standard of care; SGOT/SGPT: serum glutamic-oxalacetic transaminase/glutamic-pyruvic transaminase; SOFA: sequential Organ Failure Assessment; SPO_2_: oxygen saturation; TCZ: tocilizumab; ULN: upper limit of normal; WBC: white blood cell; WHO: World Health Organization.*
