## Supplementary file 3 for "Tocilizumab and mortality in hospitalised patients with covid-19. A systematic review comparing randomised trials with observational studies"

| Author | Bias due to confounding | Bias in selection of participants into the study | Bias in classification of interventions | Bias due to deviations from intended intervention | Bias due to missing data | Bias in measurement of outcomes | Bias in selection of the reported result | Based on Maximum Criterion |
| --- | --- | --- | --- | --- | --- | --- | --- | --- |
| ADJUSTED | | | | | | | | |
| Ruiz-Antorán et al. | 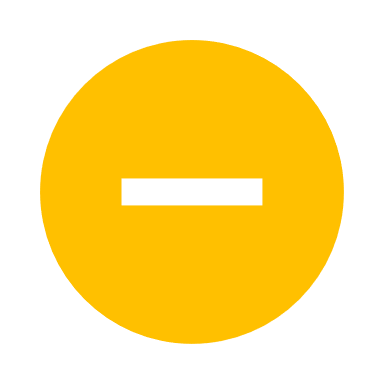 | 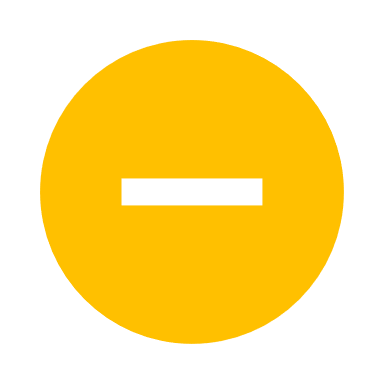 | 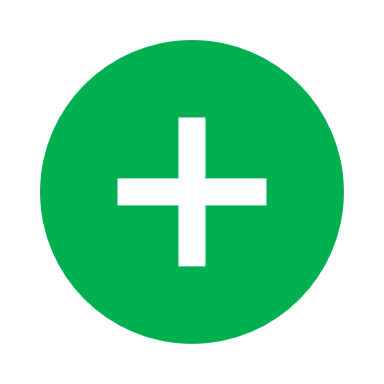 | 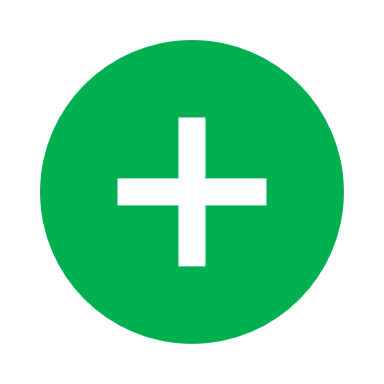 | 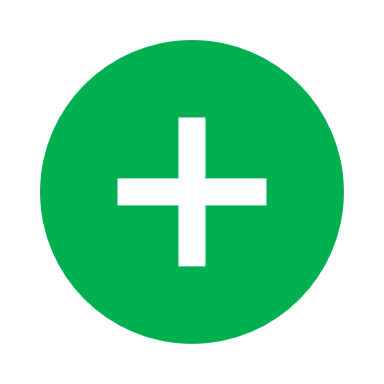 | 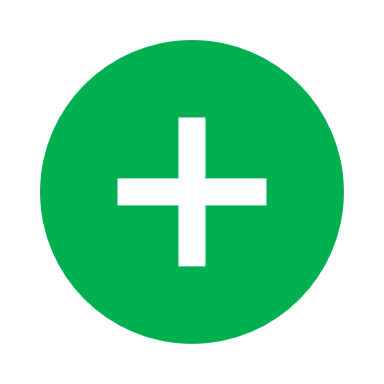 | 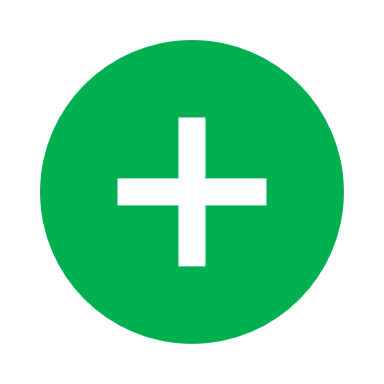 | Moderate |
| Gupta et al. | 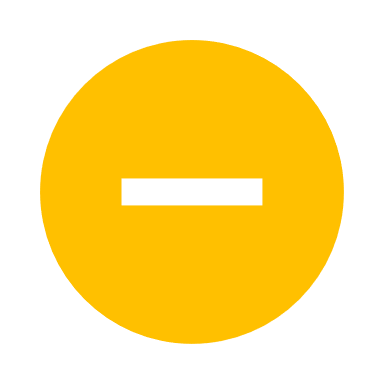 | 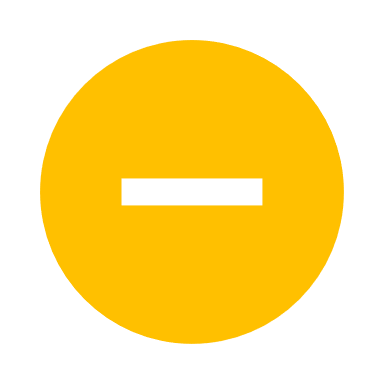 | 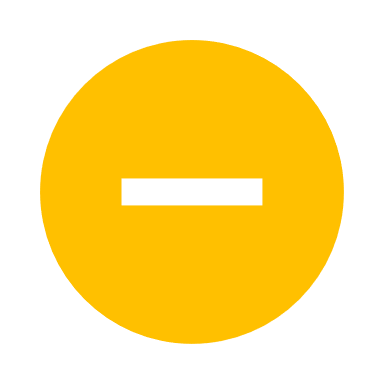 | 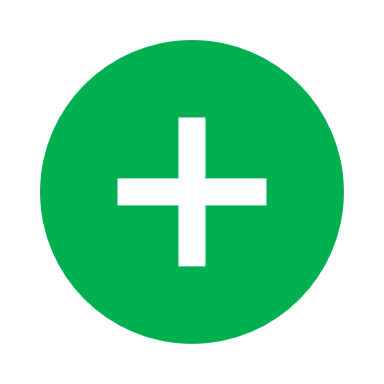 | 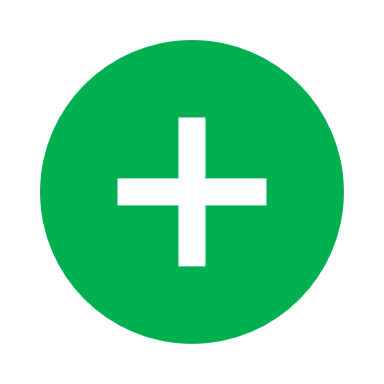 | 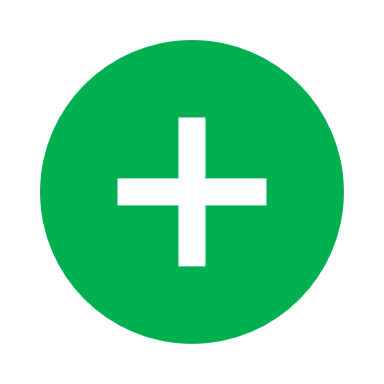 | 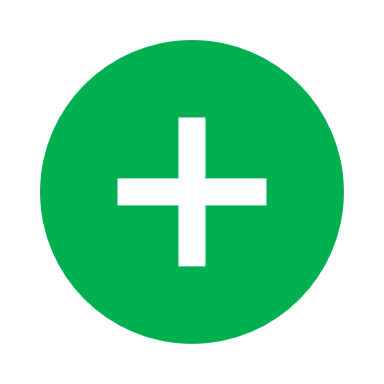 | Moderate |
| Biran et al. | 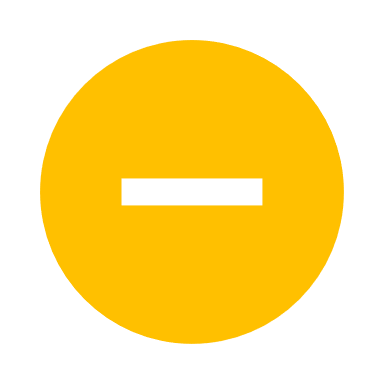 | 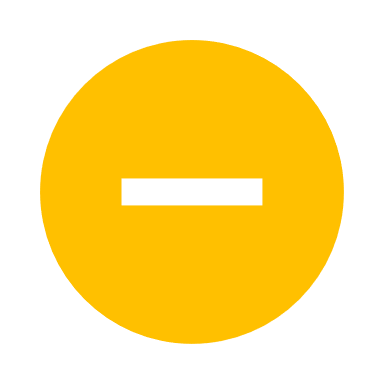 | 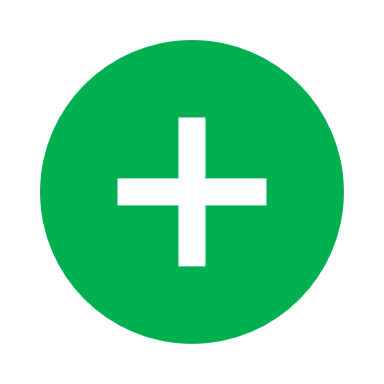 | 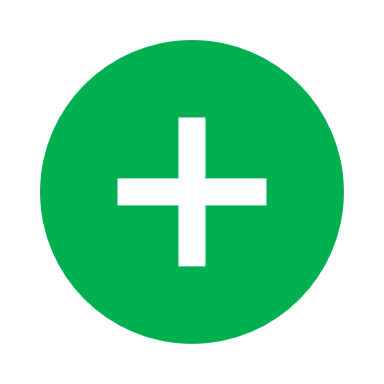 | 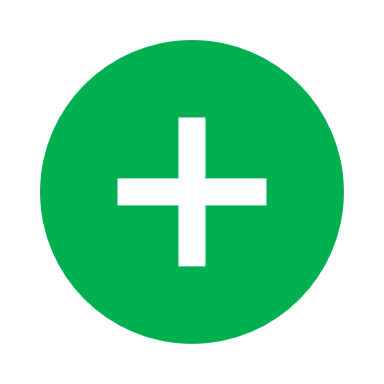 | 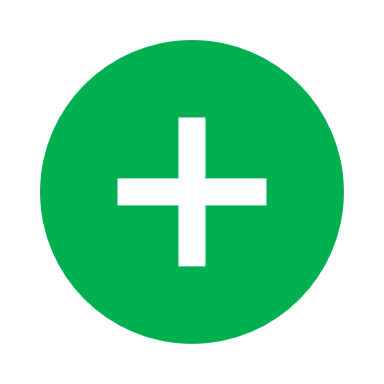 | 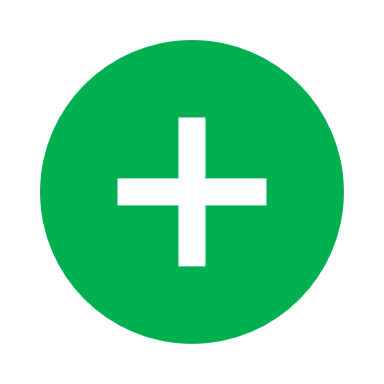 | Moderate |
| Owen et al. | 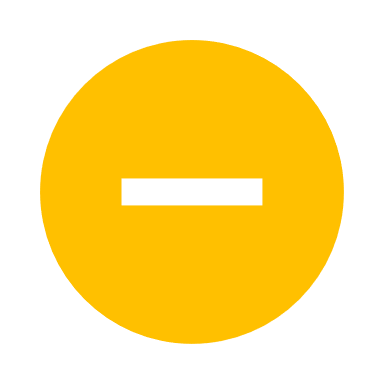 | 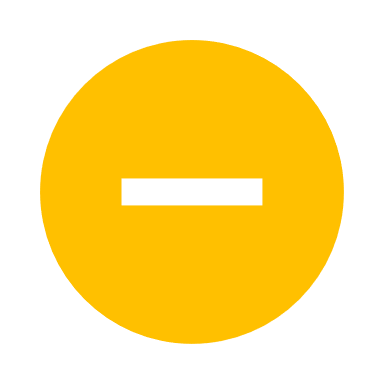 | 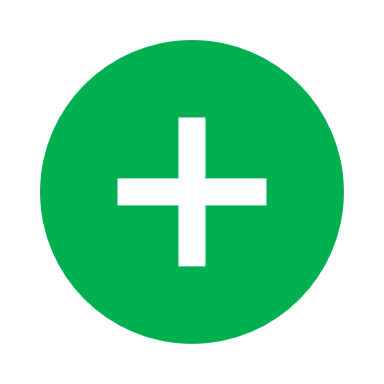 | 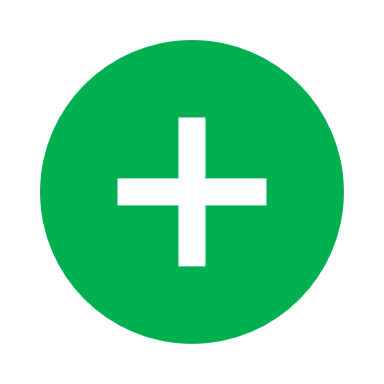 | 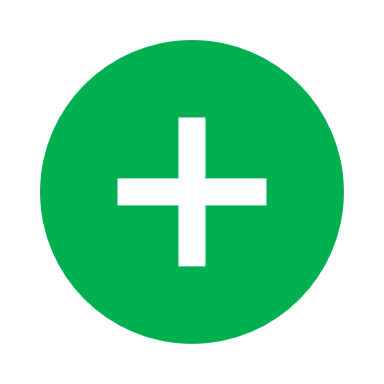 | 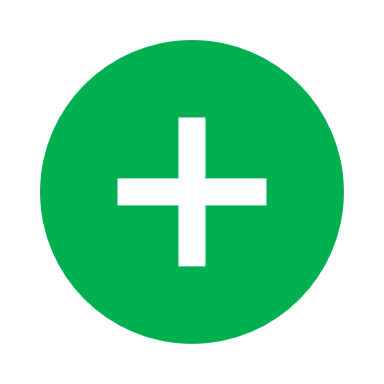 | 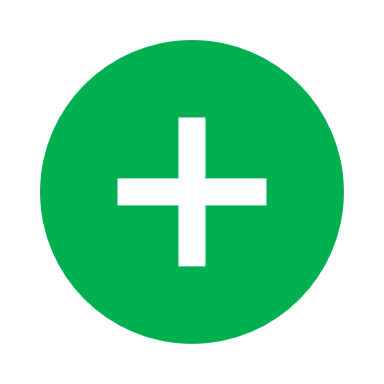 | Moderate |
| Ignatius et al. | 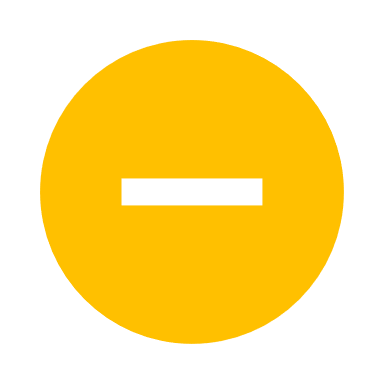 | 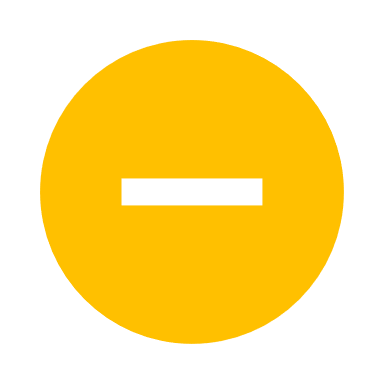 | 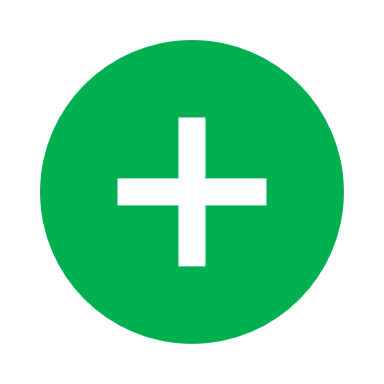 | 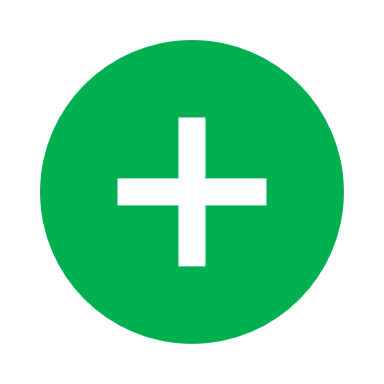 | 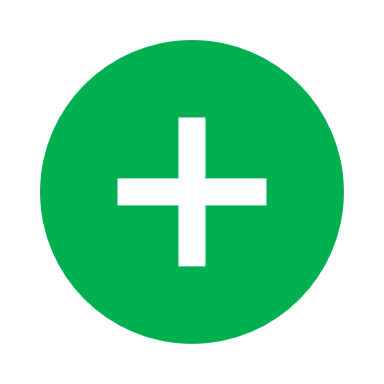 | 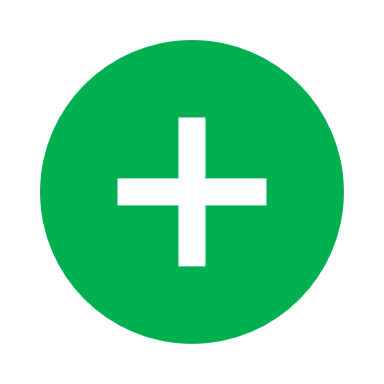 | 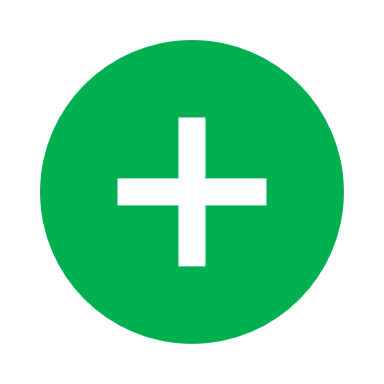 | Moderate |
| Canziani et al. | 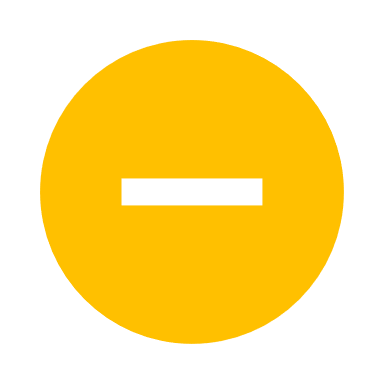 | 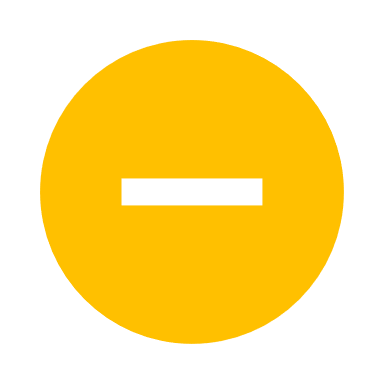 | 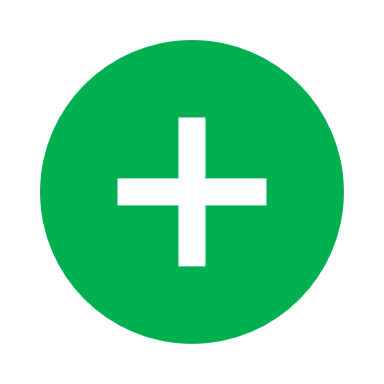 | 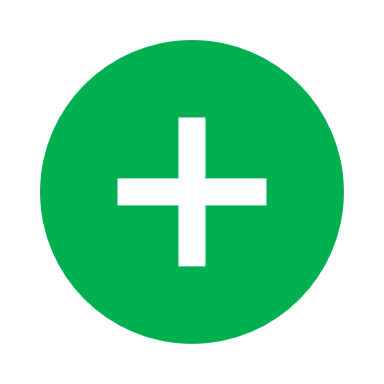 | 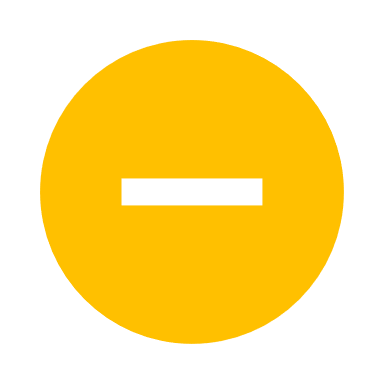 | 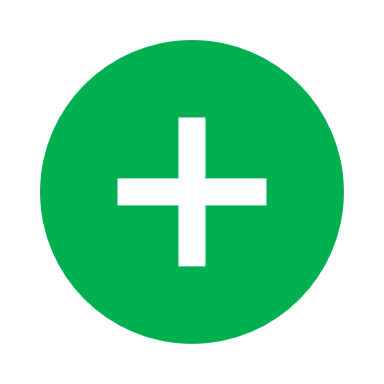 | 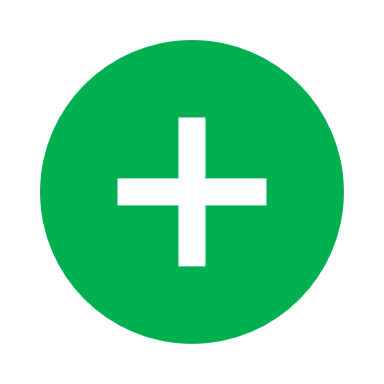 | Moderate |
| Buzón-Martín et al. | 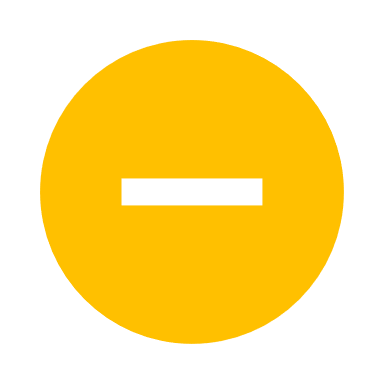 | 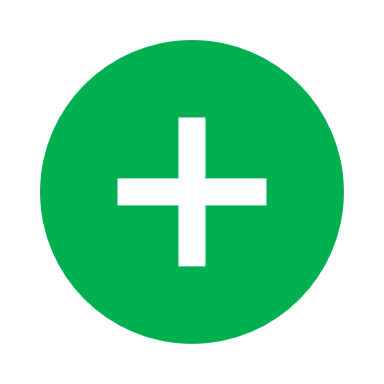 | 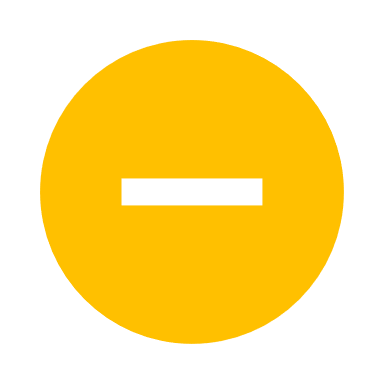 | 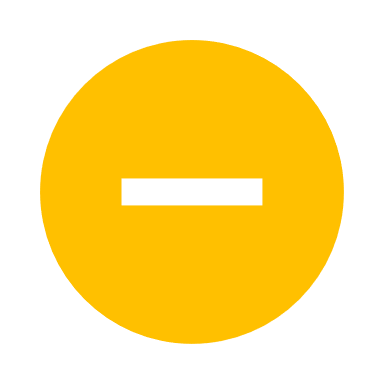 | 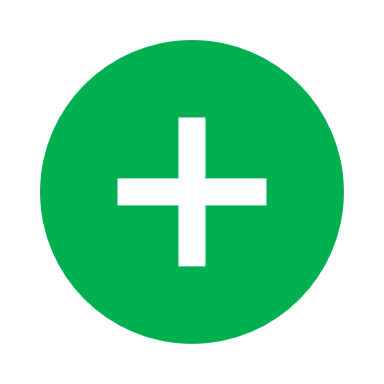 | 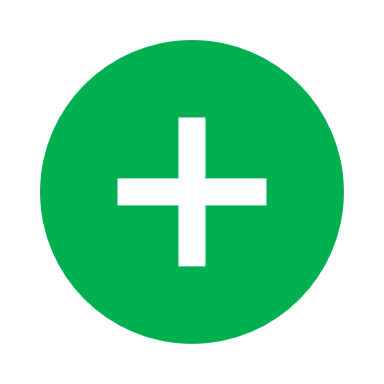 | 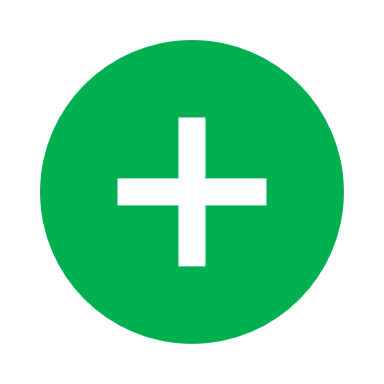 | Moderate |
| Rajendram et al. | 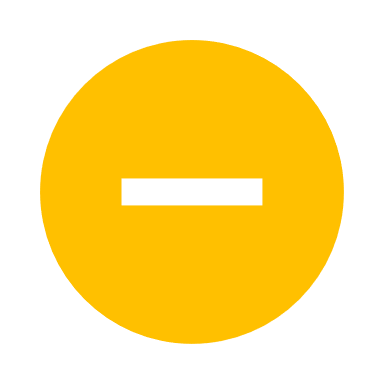 | 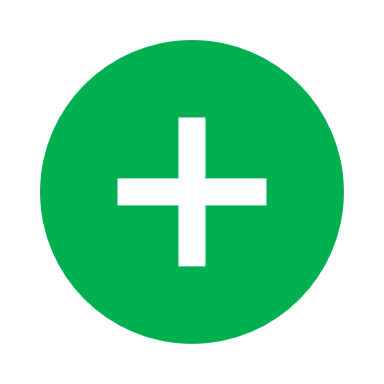 | 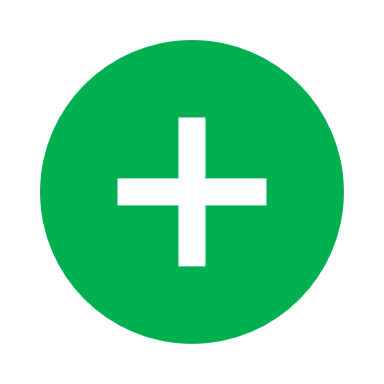 | 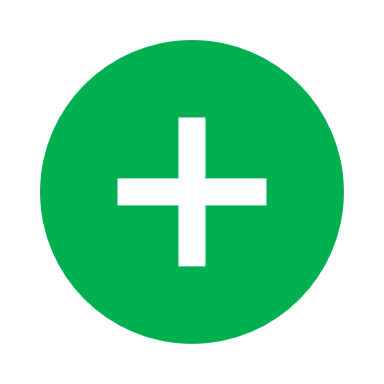 | 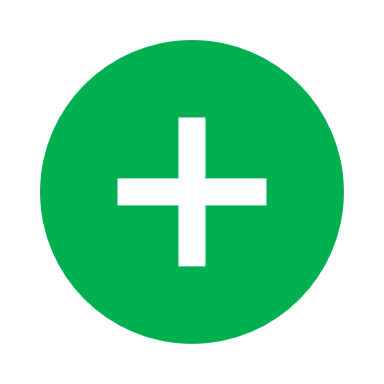 | 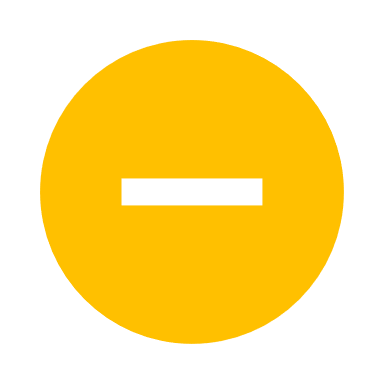 | 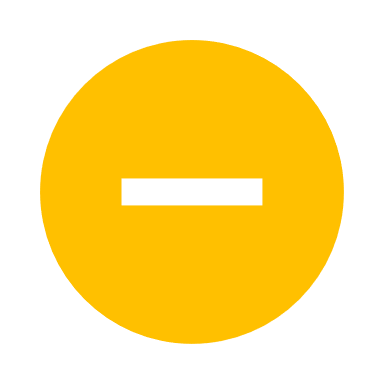 | Moderate |
| De Rossi et al. | 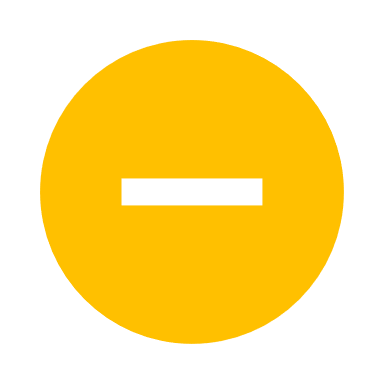 | 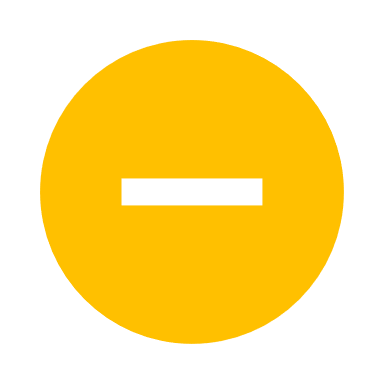 | 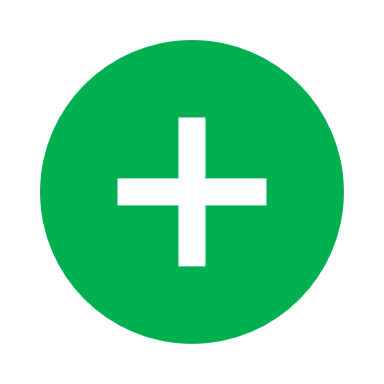 | 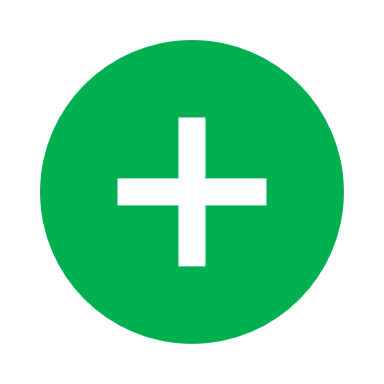 | 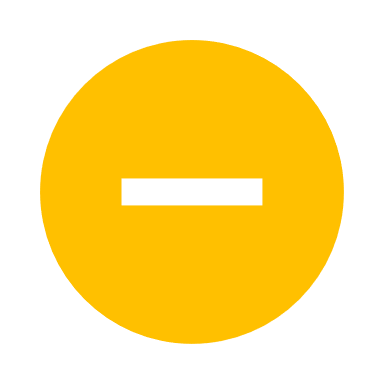 | 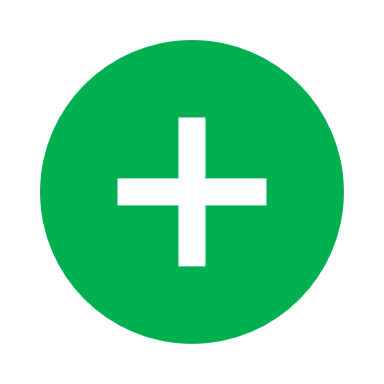 | 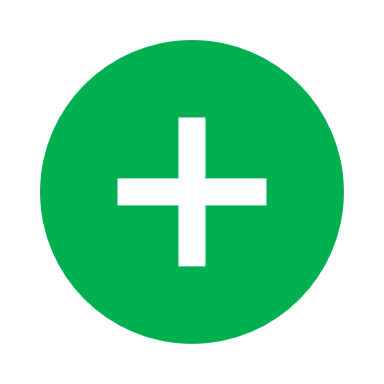 | Moderate |
| Rodríguez-Baño et al. | 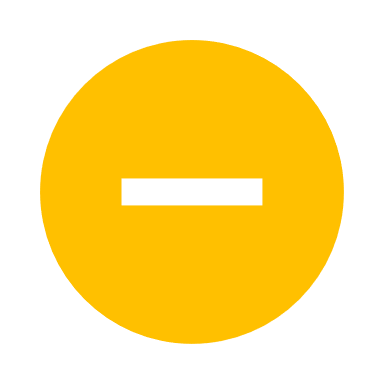 | 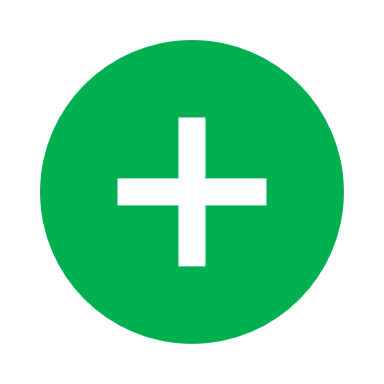 | 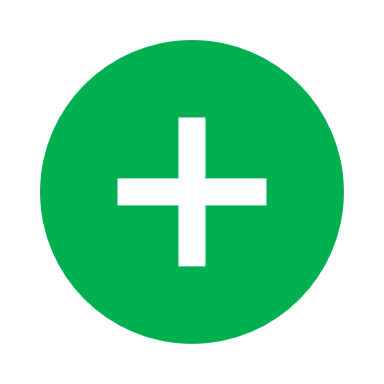 | 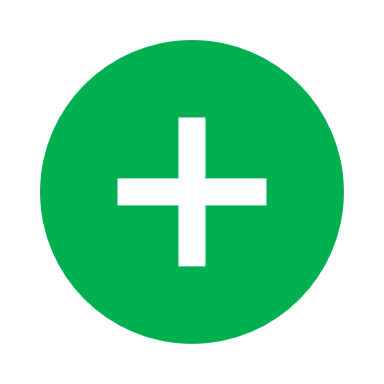 | 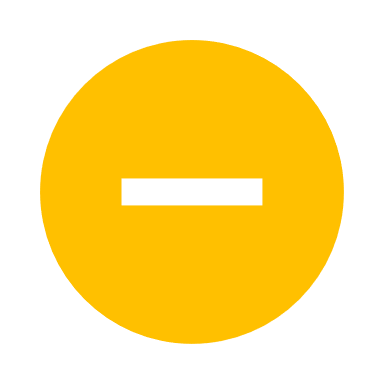 | 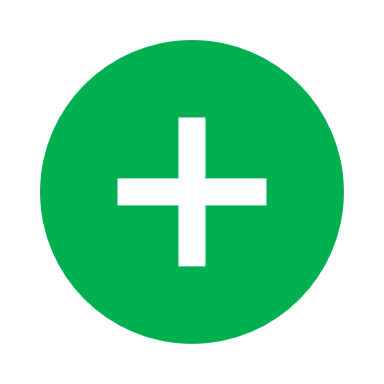 | 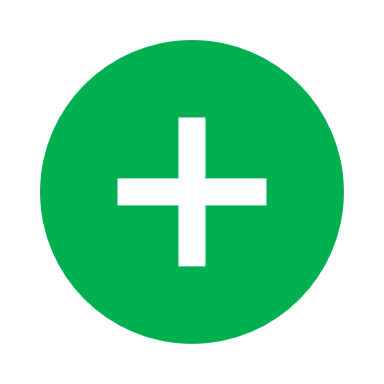 | Moderate |
| Somers et al. | 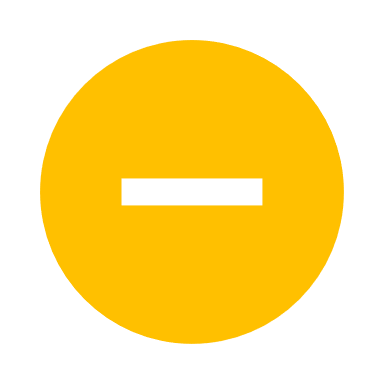 | 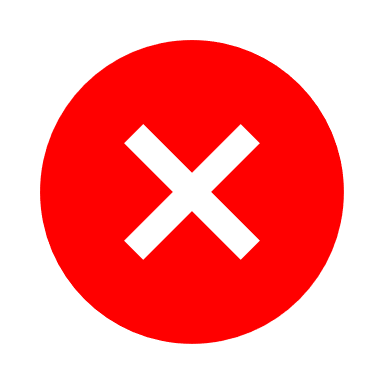 | 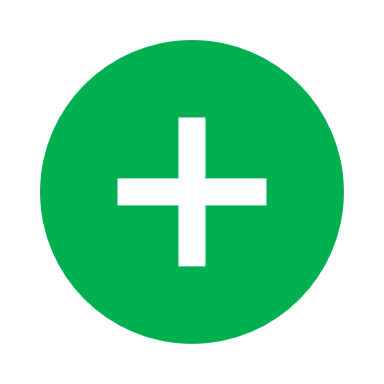 | 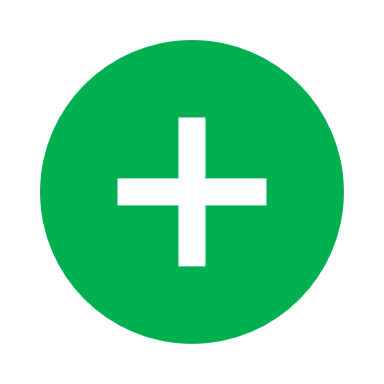 | 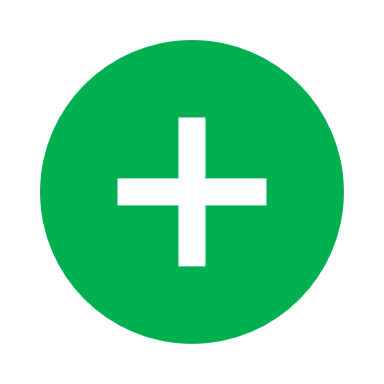 | 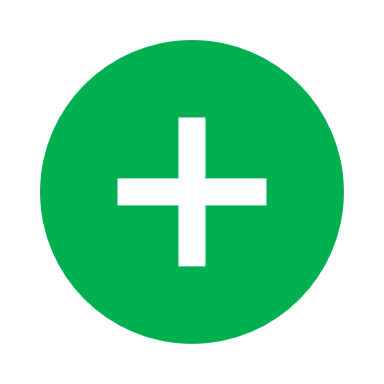 | 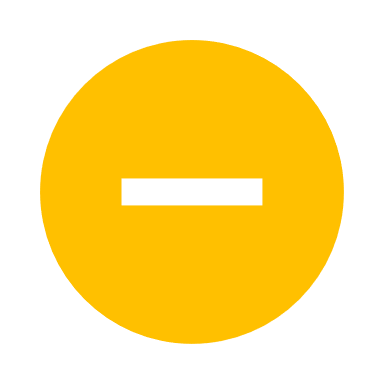 | Serious |
| Narain et al. | 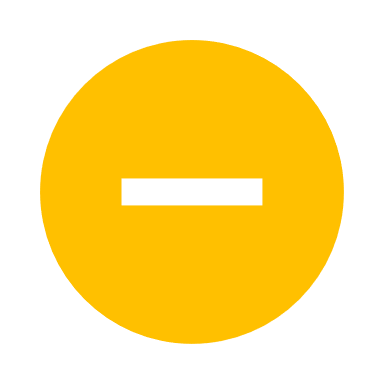 | 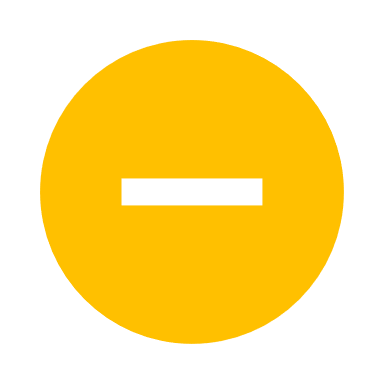 | 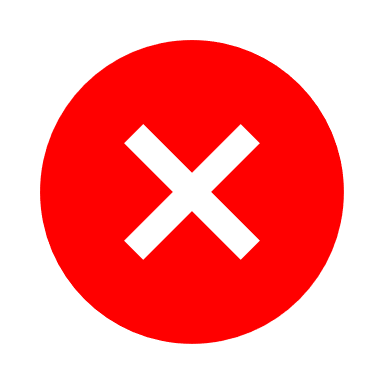 | 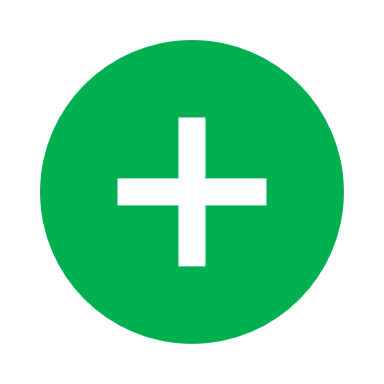 | 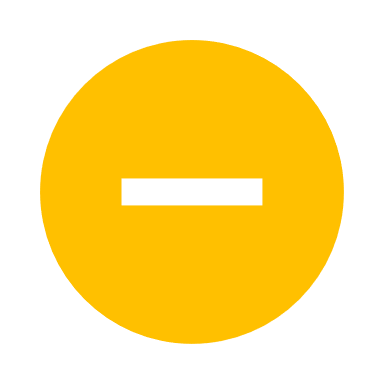 | 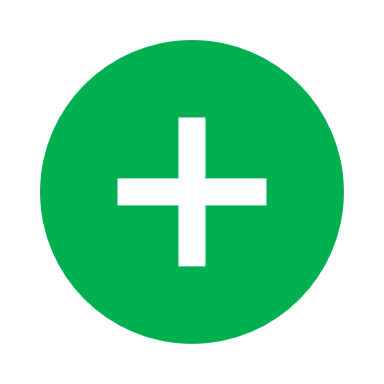 | NA | Serious |
| Tian et al. | 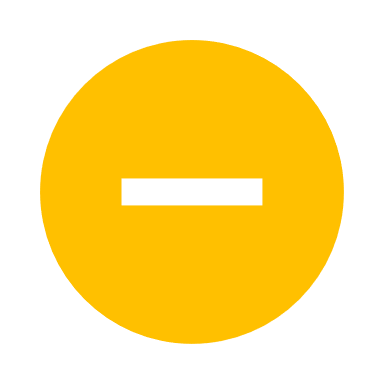 | 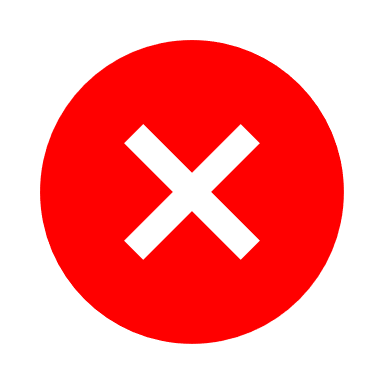 | 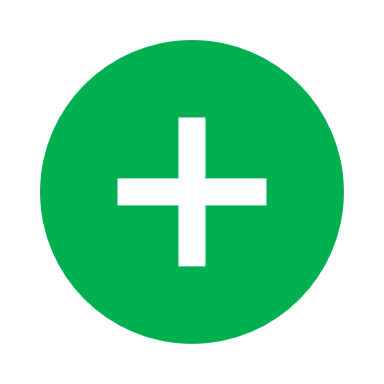 | 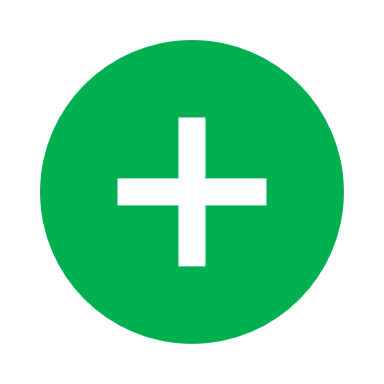 | 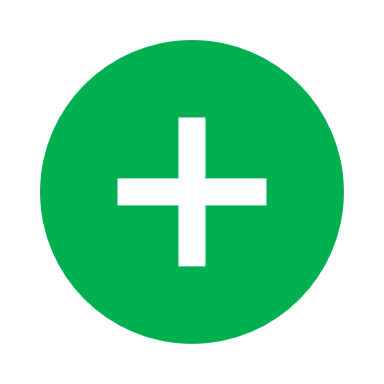 | 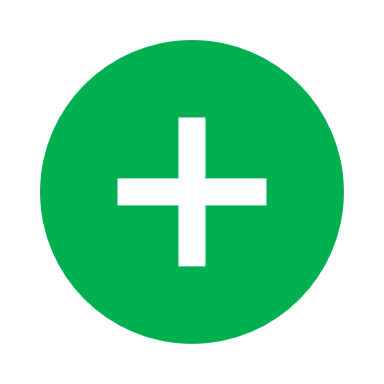 | 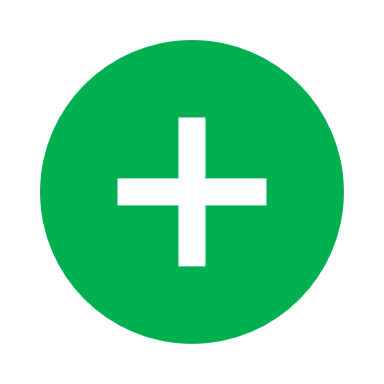 | Serious |
| Rossotti et al. | 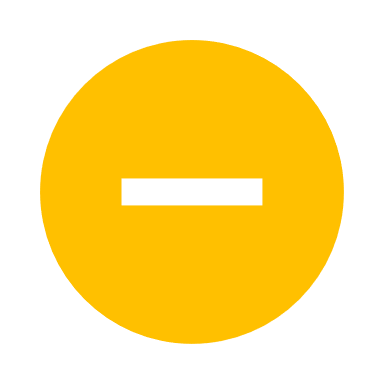 | 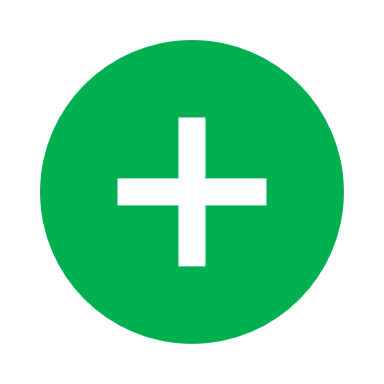 | 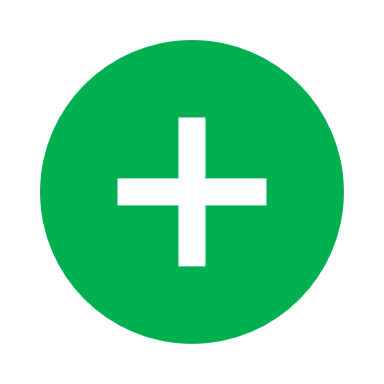 | 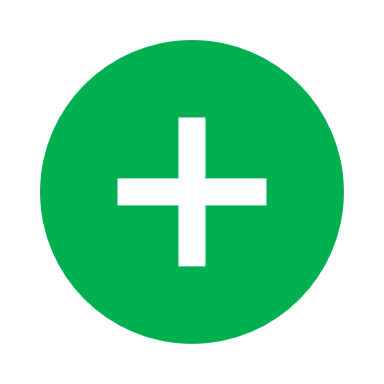 | 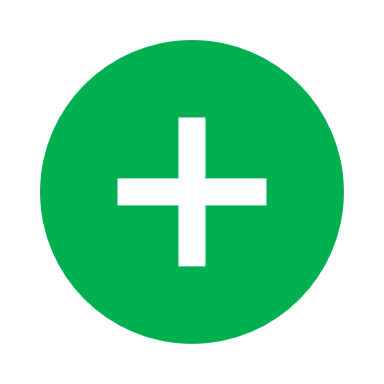 | 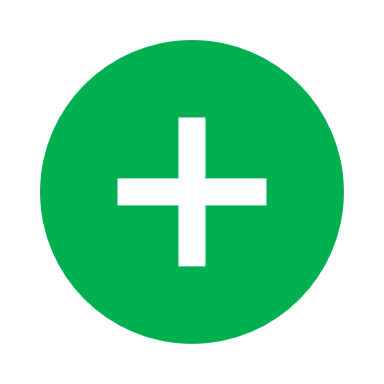 | NA | Serious |
| Rossi et al. | 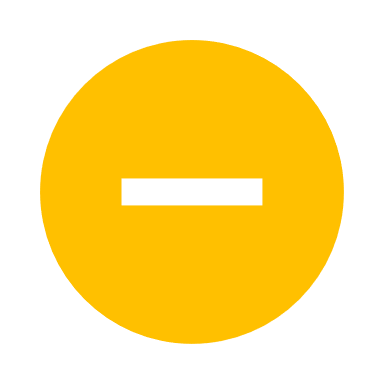 | 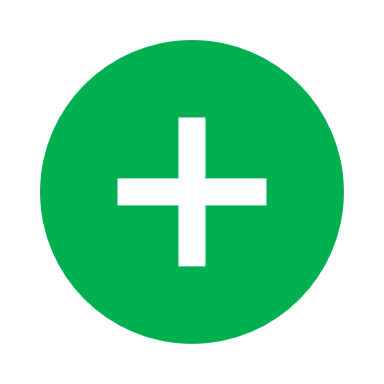 | 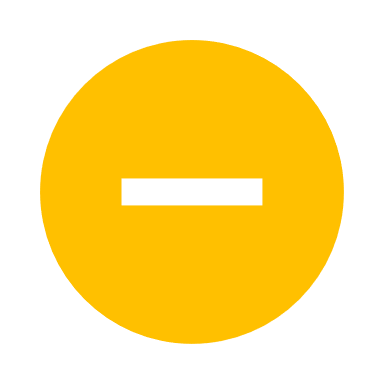 | 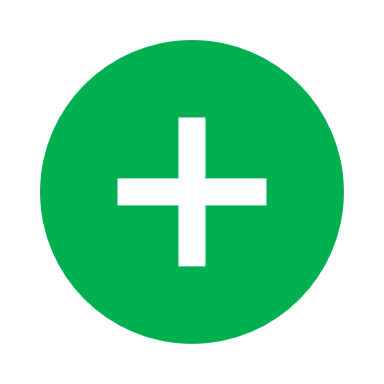 |  |  |  | Serious |
| Guisado-Vasco et al. |  |  |  |  |  |  |  | Serious |
| Menzella et al. |  |  |  |  | NA |  |  | Serious |
| Patel et al. |  |  |  |  | NA |  |  | Serious |
| Eimer et al. |  |  |  |  | NA |  |  | Moderate |
| Galván-Román et al. |  |  |  |  |  |  |  | Serious |
| Fisher et al. |  |  |  |  | NA |  |  | Serious |
| Okoh et al. |  |  |  |  | NA |  |  | Serious |
| Pereira et al. |  |  |  | NA | NA |  |  | Critical |
| UNADJUSTED | | | | | | | | |
| Campochiaro et al. |  |  |  |  | NA |  |  | Serious |
| Klopfenstein et al. |  |  |  | NA | NA |  |  | Serious |
| Masia et al. |  |  |  |  | NA |  |  | Serious |
| Vazquez Guillamet et al. |  | NA | NA | NA | NA |  |  | Critical |
| Rojas-Marte et al. |  |  |  |  |  |  |  | Critical |
| Huang et al. |  |  |  |  |  |  |  | Critical |
| Khamis et al. |  |  |  |  | NA |  |  | Critical |
| Nasa et al. |  |  |  |  |  |  |  | Critical |
| Salvati et al. |  |  |  |  | NA |  |  | Critical |
| Quartuccio et al. |  |  |  |  | NA |  |  | Critical |

- Low; - Moderate; - Serious; - Critical; NA- No information
