## Supplementary figures and images for "Tocilizumab and mortality in hospitalised patients with covid-19. A systematic review comparing randomised trials with observational studies"

### Supplementary file 4

 
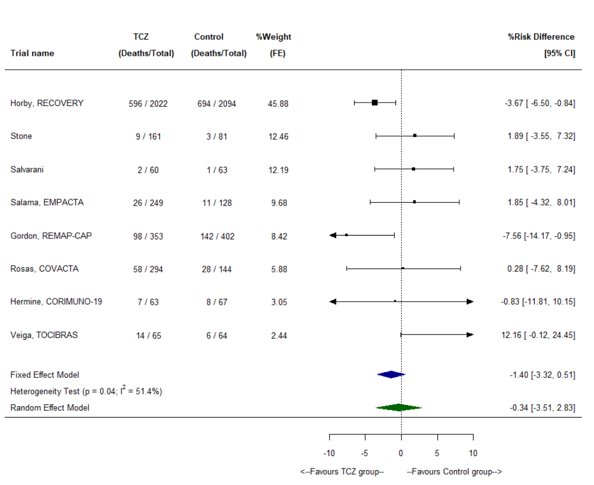
